# The MetaboHealth score enhances insulin resistance metabotyping for targeted fat loss through personalized diets: Insights from the PERSON intervention study

**DOI:** 10.1101/2024.12.18.24319249

**Authors:** Jordi Morwani-Mangnani, Fatih A. Bogaards, Alexander Umanets, Gabby B. Hul, Anouk Gijbels, Gijs H. Goossens, Joris Deelen, Marian Beekman, Lydia Afman, Ellen E. Blaak, P. Eline Slagboom

**Author notes:** Shared last authorship (Ellen E. Blaak, P. Eline Slagboom).

## Abstract

**Background:** We previously identified distinct muscle and liver insulin resistance (IR) metabotypes among middle-aged and older adults. The PERSON intervention study demonstrated beneficial effects of a low-fat, high-protein, high-fiber (LFHP) diet on the muscle IR metabotype group and of a high-monounsaturated fatty acid (HMUFA) diet on the liver IR metabotype group. We also generated a ^1^H-NMR metabolomics-based immune-metabolic health score (MetaboHealth) reflecting the risk of mortality, frailty, and cognitive decline. Here we explore its interaction with the IR metabotypes concerning (i) cardiometabolic health and (ii) body composition outcomes of the PERSON study. These studies enable development of precision nutrition strategies to reduce cardiometabolic risk in insulin resistant adults.

**Methods:** In the PERSON study, 242 individuals with overweight or obesity aged 40-75 years with insulin resistance belonging to two metabotypes-predominantly muscle or liver insulin resistant phenotypes-were randomized to follow either an isocaloric HMUFA diet or a LFHP diet for 12 weeks. The 184 participants for whom complete data was available were categorized according to the MetaboHealth score in tertiles (the higher the tertile, the poorer the immune-metabolic health). Metabolic outcomes were assessed via a 7-point oral glucose tolerance test and blood serum analyses. Body composition was assessed using dual-energy X-ray absorptiometry (DXA). Linear mixed models with estimated marginal means were used to analyze four-way interactions, exploring the relationships between MetaboHealth, metabotypes, and the two dietary interventions across the intervention period.

**Results:** Linear mixed models did not detect an interaction effect of baseline MetaboHealth tertiles, metabotypes, and diet with the primary cardiometabolic health outcomes. Significant four-way interactions were observed for the DXA outcomes android (β = 0.28, q-value = 0.003), gynoid (β = 0.27, q-value = 0.008), and total fat percentage (β = 0.17, q-value = 0.013) as well as fat mass index (β = 0.07, q-value = 0.018). In the higher MetaboHealth tertile, poorer immune-metabolic health, both dietary interventions resulted in comparable reductions in fat mass outcomes across both metabotypes. In the lower tertile reflecting healthier immune-metabolic health, participants with predominant muscle insulin resistance following the LFHP diet experienced greater android, gynoid, total fat percentage and fat mass index loss compared to those following the HMUFA, while those with liver insulin resistance showed better android and gynoid fat percentage following the HMUFA compared to the LFHP. Notably, MetaboHealth did not significantly change during the intervention.

**Conclusions:** Our findings suggest that personalized dietary strategies targeted to fat loss in insulin resistant middle-aged and older adults may become more effective when grouped by insulin resistance phenotype combined with MetaboHealth.

## INTRODUCTION

The rising prevalence of insulin resistance (IR) in aging populations poses a tremendous challenge to healthcare systems worldwide (1,2). With aging, excess adipose tissue mass tends to accumulate viscerally and ectopically, thereby contributing to an increased risk of developing IR-related diseases– the leading causes of morbidity and mortality in middle-aged and older adults (3,4). Weight management interventions have been demonstrated to be moderately successful in reducing IR (5). However, there is a large heterogeneity of response to lifestyle modifications, where not all individuals benefit (6,7). For this reason, the development of more personalized dietary strategies is essential to improve body composition and mitigate IR-related risks effectively (8). The manifestation of IR is not uniform; it can present as predominant muscle insulin resistance (MIR) or predominant liver insulin resistance (LIR), underscoring the need for personalized treatment strategies tailored to individual metabotypes (9,10).

The PERSonalized Glucose Optimization Through Nutritional Intervention (PERSON) study previously investigated health effects of dietary interventions designed for individuals with either the MIR or LIR metabotypes (11). Participants were assigned to one of two isocaloric dietary regimens: a low-fat, high-protein, high-fibre (LFHP) diet or a high monounsaturated fat (HMUFA) diet (12–14). Participants in the MIR group showed improvements in cardiometabolic outcomes, including peripheral insulin sensitivity, glucose homeostasis, serum triacylglycerol, and C-reactive protein, when following the LFHP diet. Conversely, the LIR group benefitted more from the HMUFA diet for these outcomes (15). However, both diets had similar effects on body weight and body composition, irrespective of IR metabotype. This lack of differential effects suggests a potential for further refinement of the initial IR metabotype classifications by introducing additional grouping criteria.

The recently developed blood-based ^1^H-NMR metabolomics-based MetaboHealth score provides a novel biomarker to assess individual immune-metabolic health comprehensively (16–18). The MetaboHealth score is derived from a study to predict all-cause mortality in 44,168 individuals from 12 cohorts, and is composed of 14 circulating metabolomic measures, including lipoprotein particle sizes, polyunsaturated fatty acid ratio, concentrations of histidine, leucine, valine, albumin, glucose, lactate, isoleucine, phenylalanine, acetoacetate, and glycoprotein acetyls (GlycA) - a marker of inflammation (19). Higher MetaboHealth scores indicate poorer immune-metabolic health and an increased risk for mortality, frailty, and cognitive decline (16,18,20).

This study aimed to determine whether the MetaboHealth score, as a global indicator of immune-metabolic health, can enhance the analysis of dietary intervention efficacy beyond the grouping of participants by tissue-specific IR metabotype. Specifically, we explored how baseline MetaboHealth score tertiles were associated with dietary intervention-induced changes in (i) cardiometabolic outcomes, and (ii) body composition, including metrics of fat and lean mass (15). By integrating this additional grouping, we seek to refine precision nutrition strategies.

## METHODS

### Study Design

This study was part of the PERSON study, a two-center randomized double-blind controlled trial designed to assess the effects of metabotype-specific dietary interventions on cardiometabolic health in a middle-aged and older population (FIGURE 1). A total of 242 men and women with overweight or obesity aged 40 to 75 years, with a BMI of 25 to 40 kg/m², participated in this study. Participants were screened using a 7-point oral glucose tolerance test (OGTT) to determine the hepatic insulin resistance index and muscle insulin sensitivity index. These indices were used to identify participants with predominant MIR or LIR. Eligible participants were then randomly assigned to follow either an HMUFA or a LFHP diet for 12 weeks. Participants underwent extensive characterization at baseline and the end of the 12-week intervention, as described in more detail below. The trial took place between May 2018 and November 2021 at Maastricht University Medical Center^+^ and Wageningen University & Research in the Netherlands, adhering to the principles of the Declaration of Helsinki. The study protocol received approval from the Medical Ethical Committee of MUMC^+^ (NL63768.068.17) and was registered at ClinicalTrials.gov (NCT03708419). Written informed consent was obtained from all participants.

**FIGURE 1.**
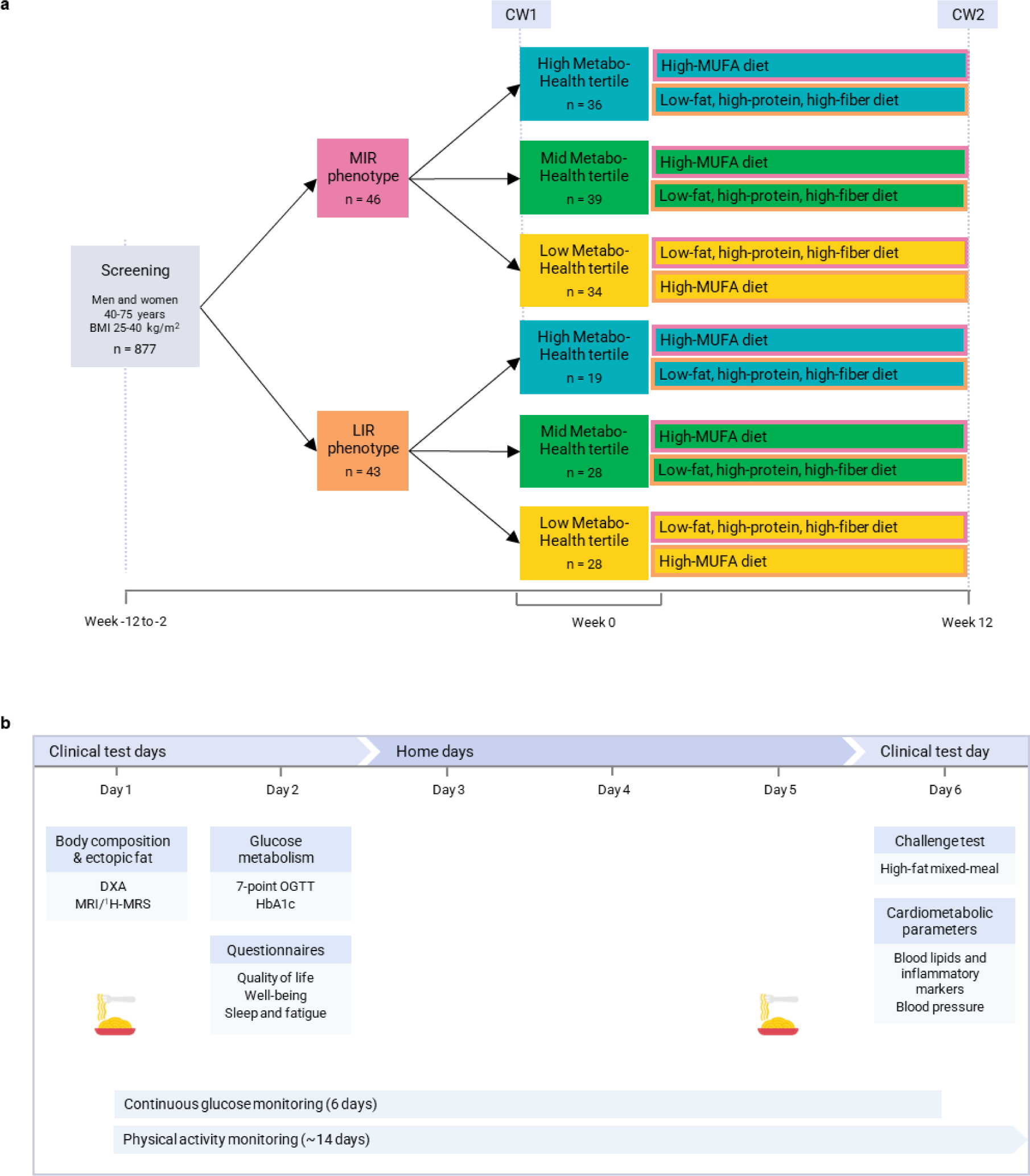
PERSON Study Design. (A) Participants were initially classified into two groups based on the predominant type of insulin resistance—muscle insulin resistance (MIR) or liver insulin resistance (LIR)—using a 7-point oral glucose tolerance test (OGTT) at screening. After classification. participants were further categorized into MetaboHealth tertiles—Low, Medium, or High—based on baseline ^1^H-NMR metabolomics measurements. Participants in all groups followed one of two dietary interventions during the study period: a high-monounsaturated fatty acid (HMUFA) diet and a low-fat. high-protein. high-fiber (LFHP) diet. The order in which participants followed each diet was randomized. allowing each phenotype to experience both diets during the intervention. (B) Clinical assessments and at-home measurements were conducted at two time points: week 0 (the start of the study) and week 12 (the end of the intervention). These assessments included comprehensive evaluations of metabolic health, physical activity, and dietary adherence. Adapted from (Trouwborst et al.. 2023)

### Assessment of Tissue-Specific Insulin Resistance (IR)

Tissue-specific insulin resistance was assessed at screening using an OGTT, where participants ingested a 75 g glucose solution. Blood samples were collected at fasting (0 minutes) and at 15, 30, 45, 60, 90, and 120 minutes post-ingestion to measure plasma glucose and insulin concentrations, from which the HIRI and MISI were calculated (11). Using tertile cutoffs for MISI and HIRI, 163 participants at screening were divided into four groups: “No MIR/LIR,” “MIR,” “LIR,” or “combined MIR/LIR.” MIR was indicated by the lowest MISI tertile, whereas LIR was indicated by the greatest HIRI tertile. Eligible participants began the intervention within 3 months of screening.

### Dietary intervention

The HMUFA diet provided 38% of energy from fat (20% MUFA, 8% PUFA, 8% SFA), 42% from carbohydrates (30% polysaccharides; 3 g/MJ fiber), and 14% from protein. The LFHP diet consisted of 28% fat (10% MUFA, 8% PUFA, 8% SFA), 42% carbohydrates (30% polysaccharides; >4 g/MJ fiber), and 24% protein. Key products that participants were provided with for the HMUFA diet included olive oil, olives, and low-fat margarine, while for the LFHP diet, these included low-fat yogurt, reduced-fat cheese, and a fiber supplement (2 g β-glucan per 6 g). Participants were instructed to consume prescribed portions daily, with alcohol limited to one glass per day. Both diets were in line with the Dutch dietary guidelines (21).

Energy intake was personalized, ranging from 6 to 13 MJ/day, based on each participant’s estimated needs, calculated from self-reported dietary intake and physical activity. Weekly counseling sessions were provided to ensure diet adherence, monitor weight stability, and address any concerns. Details of the intervention strategy have been described elsewhere (11).

### Measurements

Participants were extensively phenotyped at baseline and week 12, during a characterization week that included clinical test days and at-home data collection. Participants were instructed to refrain from alcohol and vigorous physical activity the day before and during the characterization weeks.

### 7-Point Oral Glucose Tolerance Test (OGTT)

The 7-point OGTT was conducted following the same procedures as the screening visit. Participants consumed a standardized low-fat meal the evening before the test and remained fasted until the OGTT. The primary outcome, the disposition index, was calculated as: [Matsuda index ∗ (AUC30 min insulin / AUC30 min glucose)], where AUC30 min is the area under the curve for insulin and glucose from baseline to 30 minutes. Matsuda index was calculated with the following formula: [10,000 ÷ square root of [fasting plasma glucose (mg/dL) × fasting insulin (mU/L)] × [mean glucose (mg/dL) x mean insulin (mU/L)]], using glucose and insulin values of time points 0, 30, 60, 90, and 120 min. Additional indices including HOMA-IR and HOMA-β were calculated. WHO criteria were used to define glucose status, including normal glucose tolerance (NGT), impaired fasting glucose (IFG), impaired glucose tolerance (IGT), and type 2 diabetes (T2DM).

### High-Fat Mixed-Meal Challenge Test and Biochemical Analysis

At least four days after the OGTT, participants underwent a high-fat mixed-meal challenge (HFMM) to assess postprandial glucose and lipid metabolism. Participants consumed the same low-fat macaroni meal as the OGTT on the night before the visit. After an overnight fast, participants consumed a liquid HFMM (350g, 2.8 MJ, 64% fat), and blood samples were collected at fasting (t = 0) and at 30, 60, 90, 120, 180, and 240 minutes post-consumption to measure triacylglycerol (TAG). C-reactive protein (CRP) was measured in fasting plasma using a Luminex immunoassay performed by DSM Nutritional Products (Kaiseraugst, Switzerland). In addition, metabolomics profiles of fasted plasma samples were determined using the 1H NMR metabolomics-based Nightingale Health platform (22–24).

### Body composition analysis

Body composition was assessed by dual-energy X-ray absorptiometry (DXA) (MUMC+, Discovery A, Hologic; WUR, Lunar Prodigy, GE Healthcare). These included measures such as android fat percentage of total android mass, gynoid fat percentage of total gynoid mass, total fat percentage, total lean mass percentage, fat mass index (kg/m²), lean mass index (kg/m²), and appendicular lean mass index (kg/m²) (22–24). More details on the study variables are available elsewhere (11).

### MetaboHealth Score

The MetaboHealth score is a normalized composite of 14 biomarkers—total lipids in chylomicrons and extremely large VLDL (XXL-VLDL-L), total lipids in small HDL (S-HDL-L), mean diameter for VLDL particles (VLDL-D), ratio of polyunsaturated fatty acids to total fatty acids (PUFA %), glucose, lactate, histidine, isoleucine, leucine, valine, phenylalanine, acetoacetate, albumin, and glycoprotein acetyls (GlycA)—derived from its association with all-cause mortality in 44,168 individuals across 12 European cohorts (16). It assesses cardiovascular, metabolic, and muscle health, with higher scores indicating a higher mortality risk.

### Statistical analysis

We analyzed the outcomes of the intervention by contrasting the highest and lowest baseline MetaboHealth tertiles, thus including a sample size of 117 participants with a complete dataset (FIGURE 2) (20). The primary outcomes analyzed included the cardiometabolic health outcomes, MetaboHealth score, MISI, HIRI, HOMA-IR, HOMA-B, Matsuda Index, Disposition Index, C-reactive protein (CRP), and triacylglycerides (TAG). The secondary outcomes included DXA-derived body composition metrics, such as android fat percentage, gynoid fat percentage, total fat percentage, fat mass index, lean mass index, and appendicular lean mass index. Dropouts were excluded from analyses.

**FIGURE 2.**
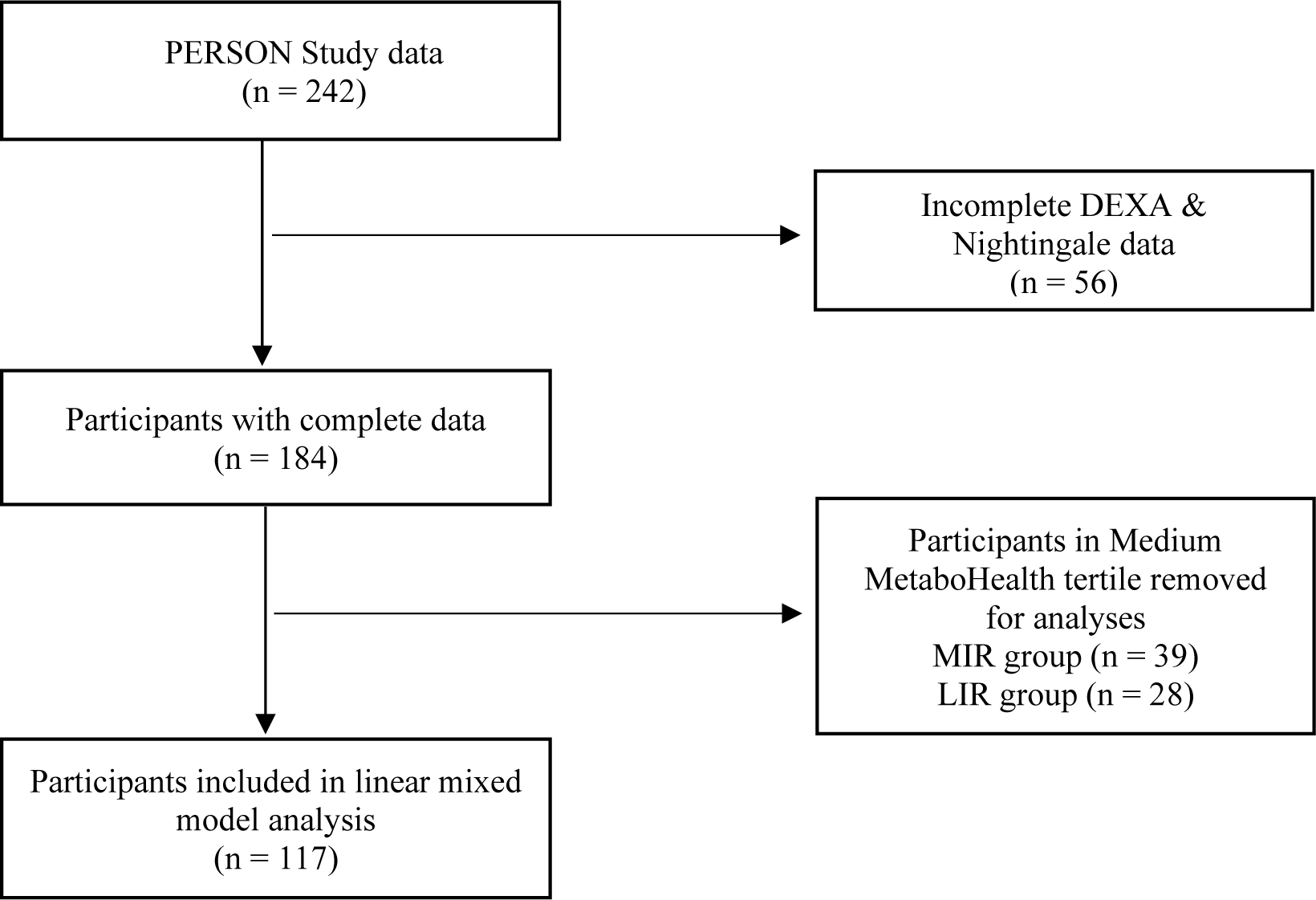
Flow diagram of the final included participants from the original PERSON Study cohort.

For the mixed-effects model analysis, traditional covariates included age, sex, BMI. Explanatory variables of interest included metabotype (MIR or LIR), diet (HMUFA or LFHP), and the MetaboHealth score tertiles defined at baseline (low versus high). The coding is detailed in TABLE S1. The model included main effects for age, sex, BMI (only for models predicting the MetaboHealth score), time, diet, metabotype, and baseline MetaboHealth tertile. Interactions included combinations of time, diet, metabotype, and MetaboHealth tertile, with a random intercept for Participant ID to account for repeated measures. The model is as follows:

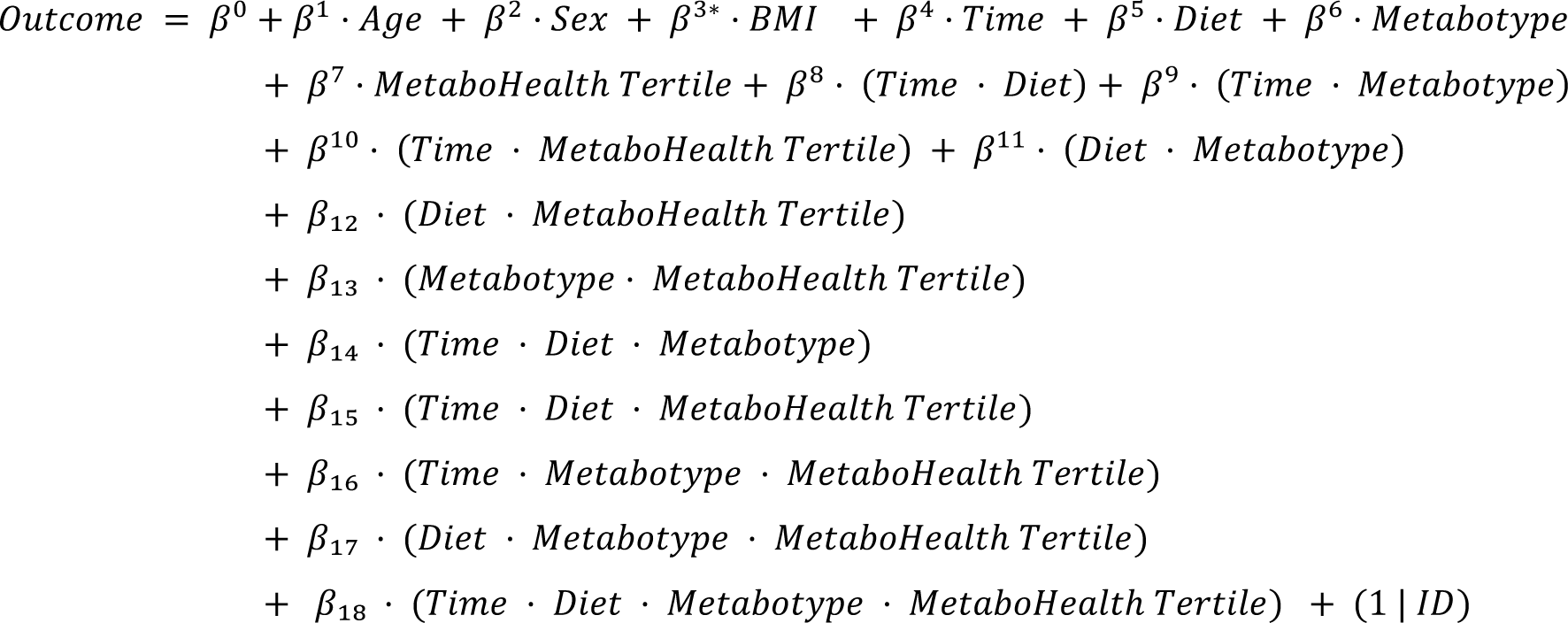

The inclusion of BMI as a covariate was restricted to models where the MetaboHealth score was the outcome, as the DXA-derived body composition measures inherently adjusted for variations in body composition. The mixed-effects models were built using the lme4 package followed by the estimation of marginal means with emmeans package (25,26). Additional packages included lme4, lmerTest, ggplot2, dyplr, tidyr in R version 4.2.3 to visualize predictive models for the four-way interactions (27–30).

## RESULTS

### Stratifying baseline metabotypes by MetaboHealth tertiles refines differences in health parameters between the groups

We investigated the baseline characteristics of the groups combining metabotype and MetaboHealth tertiles. This resulted in six groups—LIR.LowMetaboHealth (n = 28), MIR.LowMetaboHealth (n = 34), LIR.MediumMetaboHealth (n = 28), MIR.MediumMetaboHealth (n = 39), LIR.HighMetaboHealth (n = 19), and MIR.HighMetaboHealth (n = 36). We investigated the differences between these groups with cardiometabolic health and body composition outcomes. We observed differences in 13 out of 15 baseline characteristics between the groups indicating that adding the MetaboHealth to metabotype stratification further refines the group differences in key characteristics (TABLE 1).

**TABLE 1.**
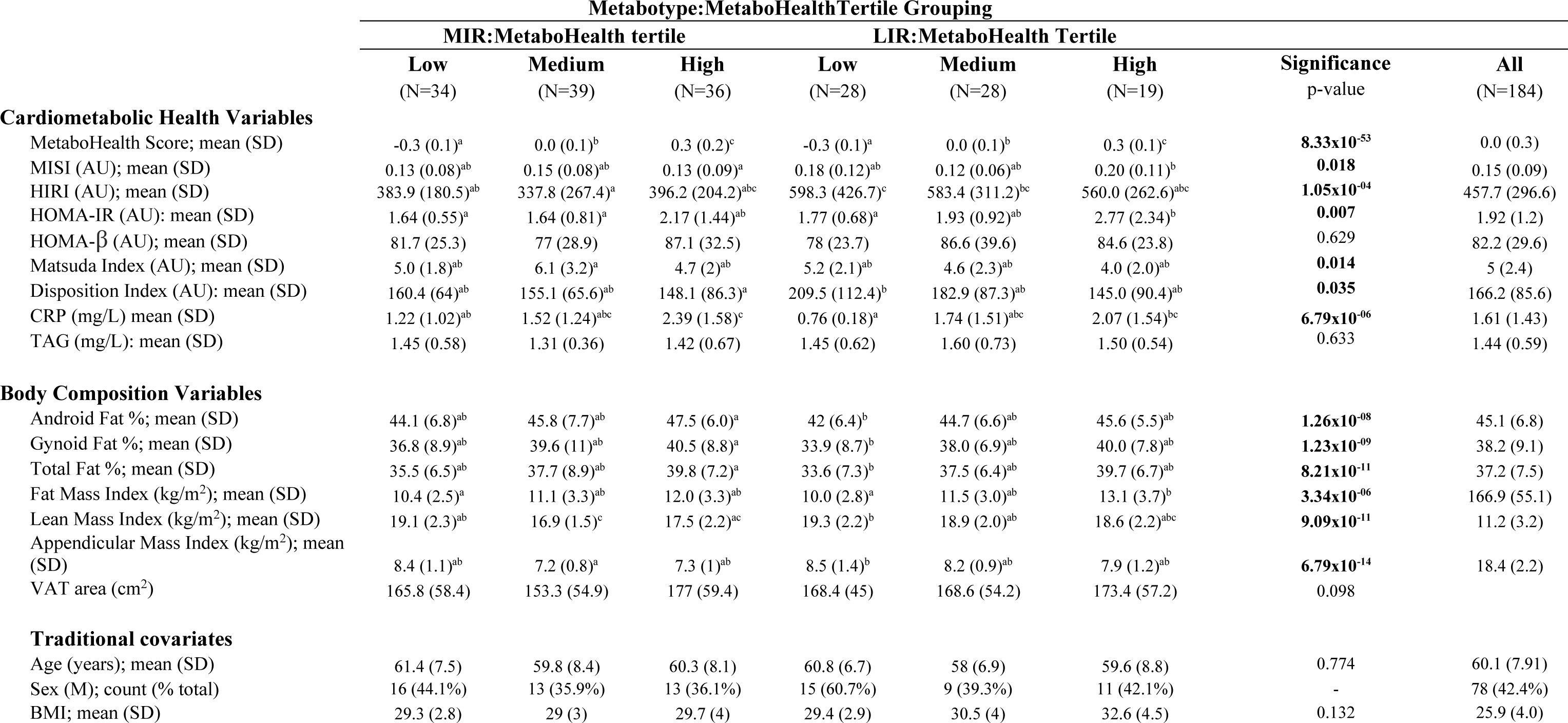
Baseline characteristics of the PERSON cohort participants selected in groups of metabotype and MetaboHealth combined. One-way ANOVA and Tukey-HSD tests were carried out to determine mean differences across six levels of Metabotype and MetaboHealth Tertiles (Low-Medium-High). The letters (a, b, c, etc.) indicate Tukey HSD groupings, where shared letters mean no significant difference, and different letters indicate significant differences (p < 0.05). CRP. C-reactive protein; HIRI. hepatic insulin resistance index; MISI. muscle insulin sensitivity index; TAG. triacylglyceride; VAT. visceral adipose tissue. n represents number of participants with a complete dataset at both baseline and endline.

Despite similar ages and BMI levels across groups (p = 0.774 and p = 0.132, respectively), sex distribution varied, with the highest percentage of males in LIR.LowMetaboHealth (60.7%) and the lowest in MIR.MediumMetaboHealth (35.9%) groups. Concerning body composition measures, individuals with predominant MIR consistently had higher android, gynoid, and total fat percentages compared to those with predominant LIR (p < 0.001 for all). The Fat Mass Index was significantly higher in both MIR.HighMetaboHealth and LIR.HighMetaboHealth groups compared to the other groups (p = 3.34×10^-06^), while lean mass indices were lower in MIR.HighMetaboHealth tertiles (p = 9.09×10^-11^). Moreover, individuals in the LIR.HighMetaboHealth and MIR.HighMetaboHealth tertiles had higher CRP levels compared to those in the lower tertiles (p = 6.79×10^-06^), demonstrating that the MetaboHealth score reflects overall immune-metabolic health.

### In response to the intervention, the refined IR metabotype-MetaboHealth combined phenotypes reveal differences in fat loss, but not cardiometabolic health outcomes

For the primary outcomes, we did not find any significant four-way interactions for MetaboHealth score, Disposition index, MISI, HIRI, HOMA-IR, HOMA-β, Matsuda index, and CRP were not significant (TABLES S3).

Next, we analyzed whether grouping MetaboHealth tertiles refined response differences between the two diets for MIR and LIR metabotypes concerning body composition changes as outcome. A four-way interaction model was used to investigate differences in response based on the six groups. Significant four-way interactions between baseline metabotype, and MetaboHealth tertile, diet, and time emerged, particularly in fat percentage outcomes. TABLE 2 presents regression coefficients for all terms, with particular focus on the three- and four-way interaction of MIR and LIR metabotypes with the highest and lowest MetaboHealth tertiles across both HMUFA and LFHP diets. Individuals with the MIR metabotype following the LFHP diet had significant reductions in android fat (β = −0.71, q = 4.66×10^-14^), gynoid fat (β = −0.68, q = 0.002), and total fat percentages (β = −0.46, q = 0.001) and fat mass index (β = −0.19, q = 0.002). Similarly, MIR individuals in the high MetaboHealth tertile at baseline following the LFHP diet experienced a reduction in total fat percentage (β = −0.11, q = 0.048) upon the intervention. These MIRhigh Metabohealth participants showed a decrease in android fat percentage across the intervention regardless of diet (β = −0.16 q = 0.019). The significant four-way interaction between time, diet, metabotype, and MetaboHealth tertile revealed differential intervention effects on android, gynoid, and total fat percentages and fat mass index (β = 0.28, q = 0.003; β = 0.28, q = 0.008, β = 0.17, q = 0.013, and β = 0.07, q = 0.008, respectively). Changes in lean mass index, appendicular lean mass index, and VAT area following the intervention were not significantly different between groups (TABLES S2).

**TABLE 2.** Effect sizes showing the association between the baseline combined metabotype-MetaboHealth tertile groups and body composition changes in response to the intervention. Regression coefficients of mixed linear models exploring the four-way interaction of MIR and LIR metabotypes with Highest and Lowest MH tertiles and two diets across the intervention. FDR-adjusted p-values are presented below.

| Model Variable | Android Fat % |  | Gynoid Fat % |  | Total Fat % |  | Fat Mass Index (kg/m<sup>2</sup>) |  |
| --- | --- | --- | --- | --- | --- | --- | --- | --- |
|  | Estimate | Adjusted p-value | Estimate | Adjusted p-value | Estimate | Adjusted p-value | Estimate | Adjusted p-value |
| (Intercept) | 47.05 | **4.66x10<sup>-14</sup>** | 43.89 | **9.11x10<sup>-15</sup>** | 40.27 | **5.09x10<sup>-16</sup>** | 13.48 | **1.24x10<sup>-07</sup>** |
| Age | -0.09 | 0.518 | -0.07 | 0.580 | -0.06 | 0.580 | -0.05 | 0.271 |
| Sex(Male) | -4.21 | **0.006** | -13.78 | **1.33x10<sup>-20</sup>** | -10.78 | **3.28x10<sup>-18</sup>** | -3.35 | **6.49x10<sup>-08</sup>** |
| Time | -0.27 | 0.021 | -0.22 | 0.098 | -0.21 | **0.008** | -0.09 | **0.013** |
| diet(LFHP) | 2.45 | 0.731 | 0.87 | 0.885 | 2.50 | 0.612 | 1.07 | 0.722 |
| Metabotype(MIR) | 0.85 | 0.885 | 1.19 | 0.841 | 2.46 | 0.610 | 1.67 | 0.530 |
| MetaboHealthTertile.High | 1.88 | 0.314 | 1.58 | 0.371 | 2.62 | **0.035** | 1.57 | **0.019** |
| Time:dietLFHP | 0.19 | 0.287 | 0.38 | **0.023** | 0.26 | 0.019 | 0.10 | **0.037** |
| Time:MetabotypeMIR | 0.43 | **0.003** | 0.37 | **0.021** | 0.25 | 0.018 | 0.09 | **0.040** |
| dietLFHP:MetabotypeMIR | 1.38 | 0.884 | 0.31 | 0.971 | -3.99 | 0.549 | -2.30 | 0.535 |
| Time:MHTertileHigh | 0.05 | 0.510 | 0.03 | 0.669 | 0.03 | 0.518 | 0.01 | 0.702 |
| dietLFHP:MHTertileHigh | -0.99 | 0.758 | 0.62 | 0.840 | -1.26 | 0.610 | -0.80 | 0.549 |
| MetabotypeMIR:MHTertileHigh | 0.39 | 0.885 | -0.06 | 0.976 | -1.19 | 0.580 | -1.22 | 0.204 |
| Time:dietLFHP:MetabotypeMIR | -0.71 | **3.41x10<sup>-04</sup>** | -0.68 | **0.002** | -0.46 | **0.001** | -0.19 | **0.002** |
| Time:dietLFHP:MHTertileHigh | -0.12 | 0.170 | -0.16 | 0.058 | -0.11 | **0.048** | -0.04 | 0.098 |
| Time:MetabotypeMIR:MHTertileHigh | -0.16 | **0.019** | -0.12 | 0.106 | -0.08 | 0.098 | -0.03 | 0.204 |
| dietLFHP:MetabotypeMIR:MHTertileHigh | -0.69 | 0.879 | -1.18 | 0.750 | 1.81 | 0.564 | 1.31 | 0.400 |
| Time:dietLFHP:MetabotypeMIR:MHTertileHigh | 0.28 | **0.003** | 0.28 | **0.008** | 0.17 | **0.013** | 0.07 | **0.018** |

### Visualization of the Four-Way Interaction effects on metabotypes and diet reveals further differences in android and aynoid fat percentage outcomes across low and high MetaboHealth tertiles

The estimated marginal means were used to visualize the significant four-way interaction between baseline Metabotype and MetaboHealth tertile (contrasting low versus high), applied at baseline, Diet, and Time concerning decreases in android, gynoid, and total fat percentages and fat mass index across the intervention (FIGURES 3-6). Additional graphs for other body composition and metabolic outcomes that did not demonstrate significant four-way interactions are presented in FIGURES S1-12, with all EMM values detailed in TABLES S4-5.

**FIGURE 3.**
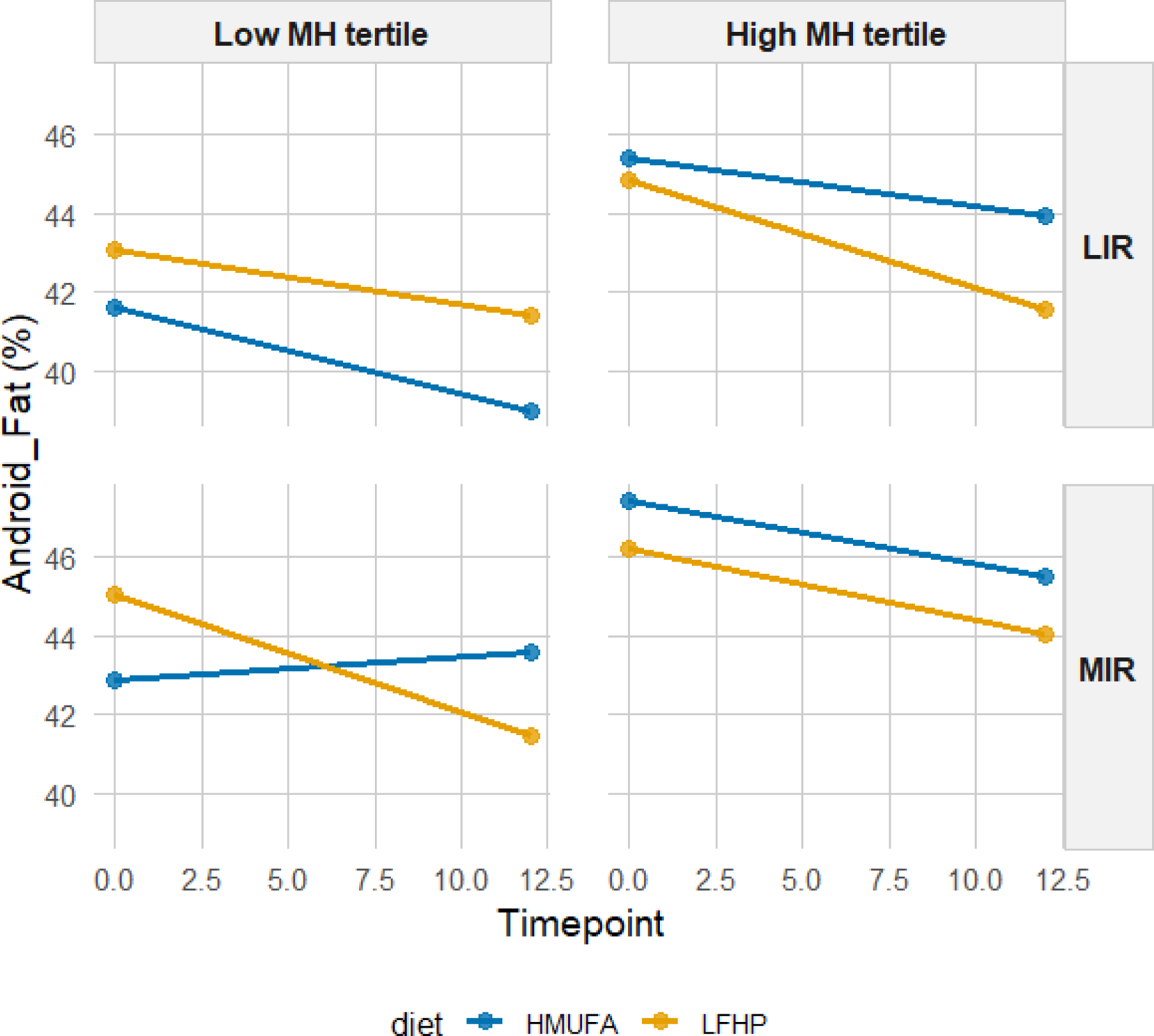
Estimated marginal means for android fat percentage by IR metabotype and MH tertiles across diets. A 2×2 matrix shows estimated total fat percentage grouped by IR metabotype (MIR vs. LIR) and MH tertiles (high vs. low). The low-fat. high-protein. high-fiber (LFHP) diet is in yellow and the high-monounsaturated fat (HMUFA) diet in blue. The range at each point represents the upper and lower confidence intervals. LIR.Low (n=28). LIR.High (n=19). MIR.Low (n=34). MIR.High (n=36).

**FIGURE 4.**
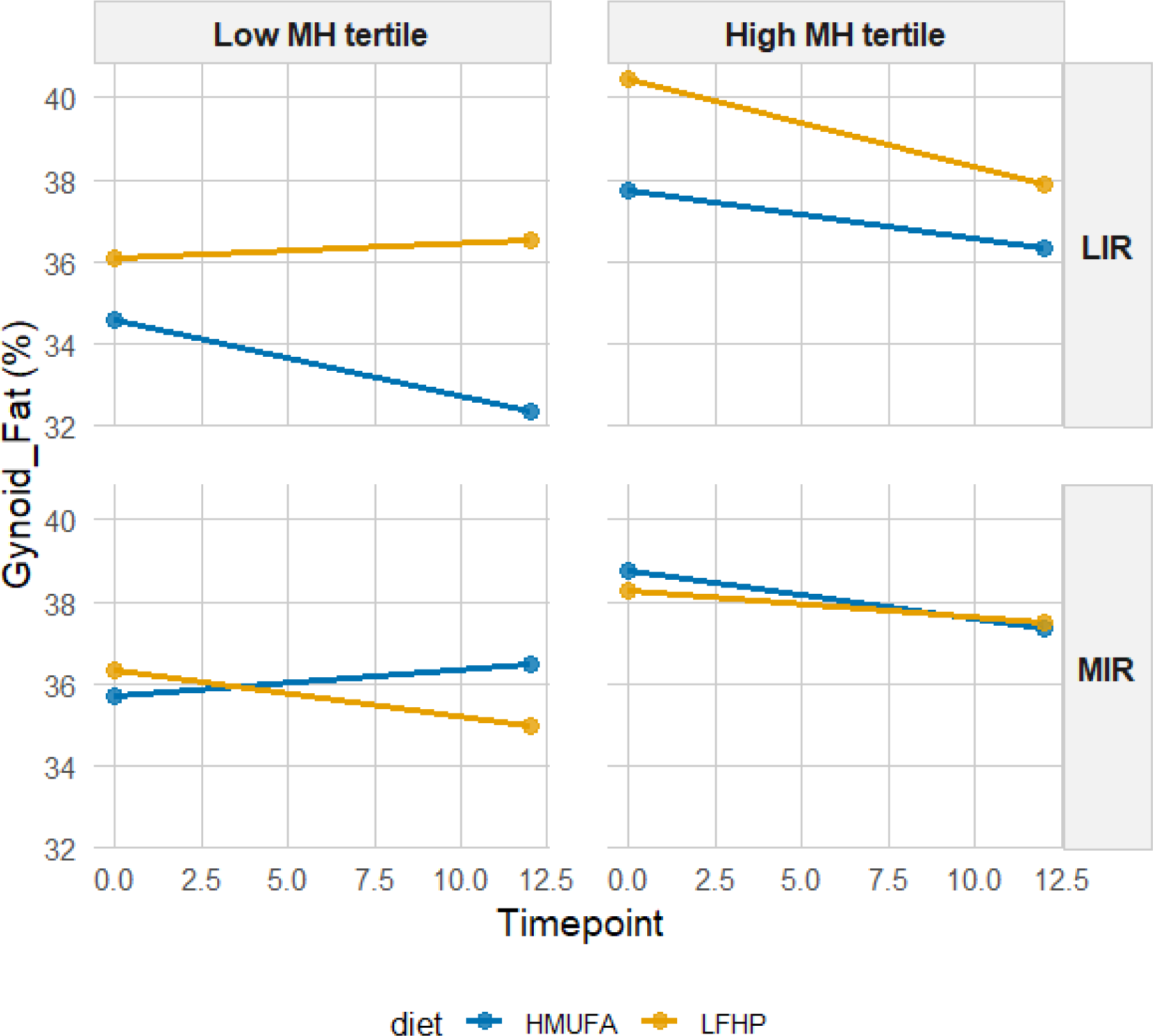
Estimated marginal means for gynoid fat percentage by IR metabotype and MH tertiles across diets. A 2×2 matrix shows estimated total fat percentage grouped by IR metabotype (MIR vs. LIR) and MH tertiles (high vs. low). The low-fat. high-protein. high-fiber (LFHP) diet is in yellow and the high-monounsaturated fat (HMUFA) diet in blue. The range at each point represents the upper and lower confidence intervals. LIR.Low (n=28). LIR.High (n=19). MIR.Low (n=34). MIR.High (n=36).

**FIGURE 5.**
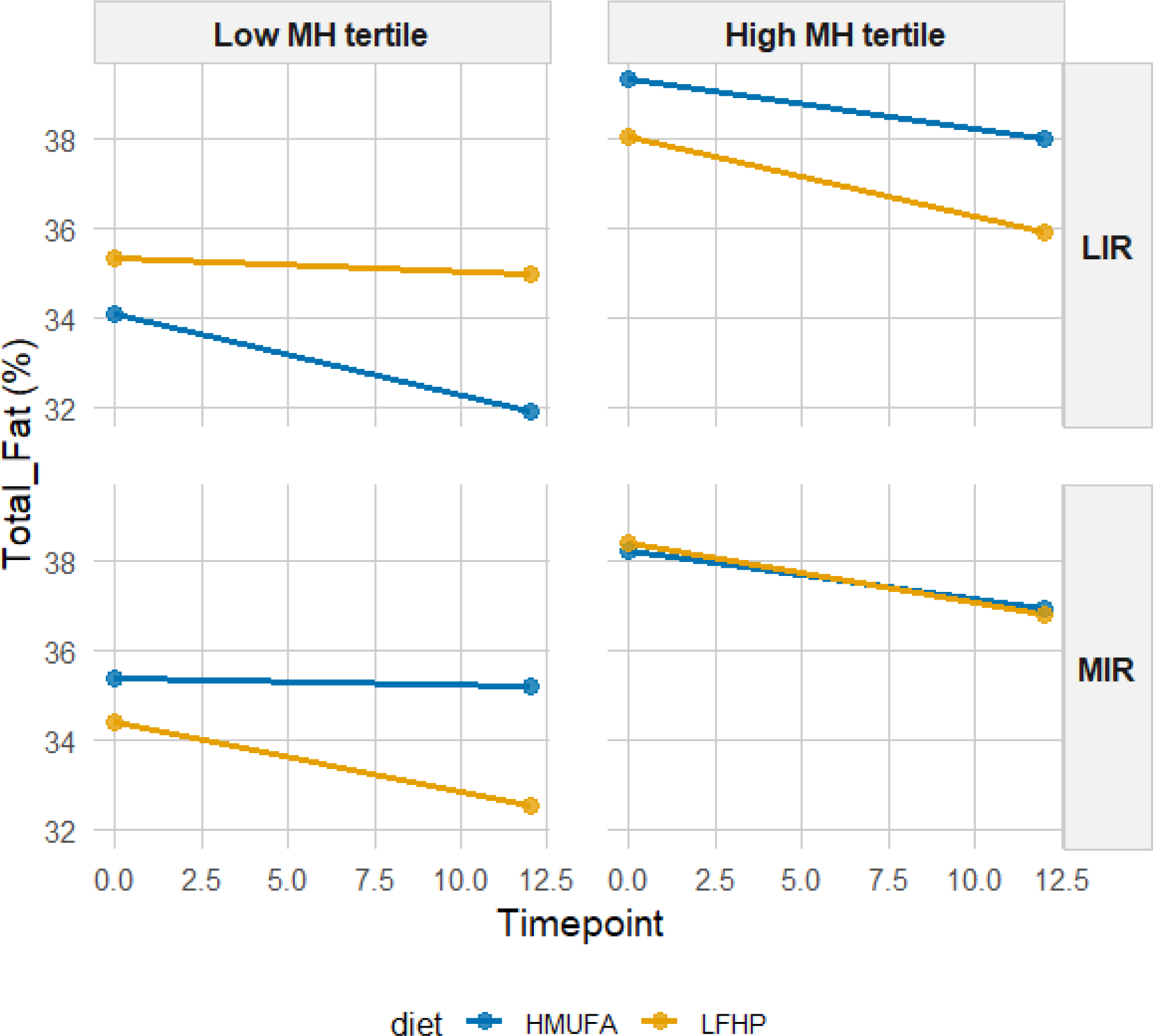
Estimated marginal means for total fat percentage by IR metabotype and MH tertiles across diets. A 2×2 matrix shows estimated total fat percentage grouped by IR metabotype (MIR vs. LIR) and MH tertiles (high vs. low). The low-fat. high-protein. high-fiber (LFHP) diet is in yellow and the high-monounsaturated fat (HMUFA) diet in blue. LIR.Low (n=28). LIR.High (n=19). MIR.Low (n=34). MIR.High (n=36).

**FIGURE 6.**
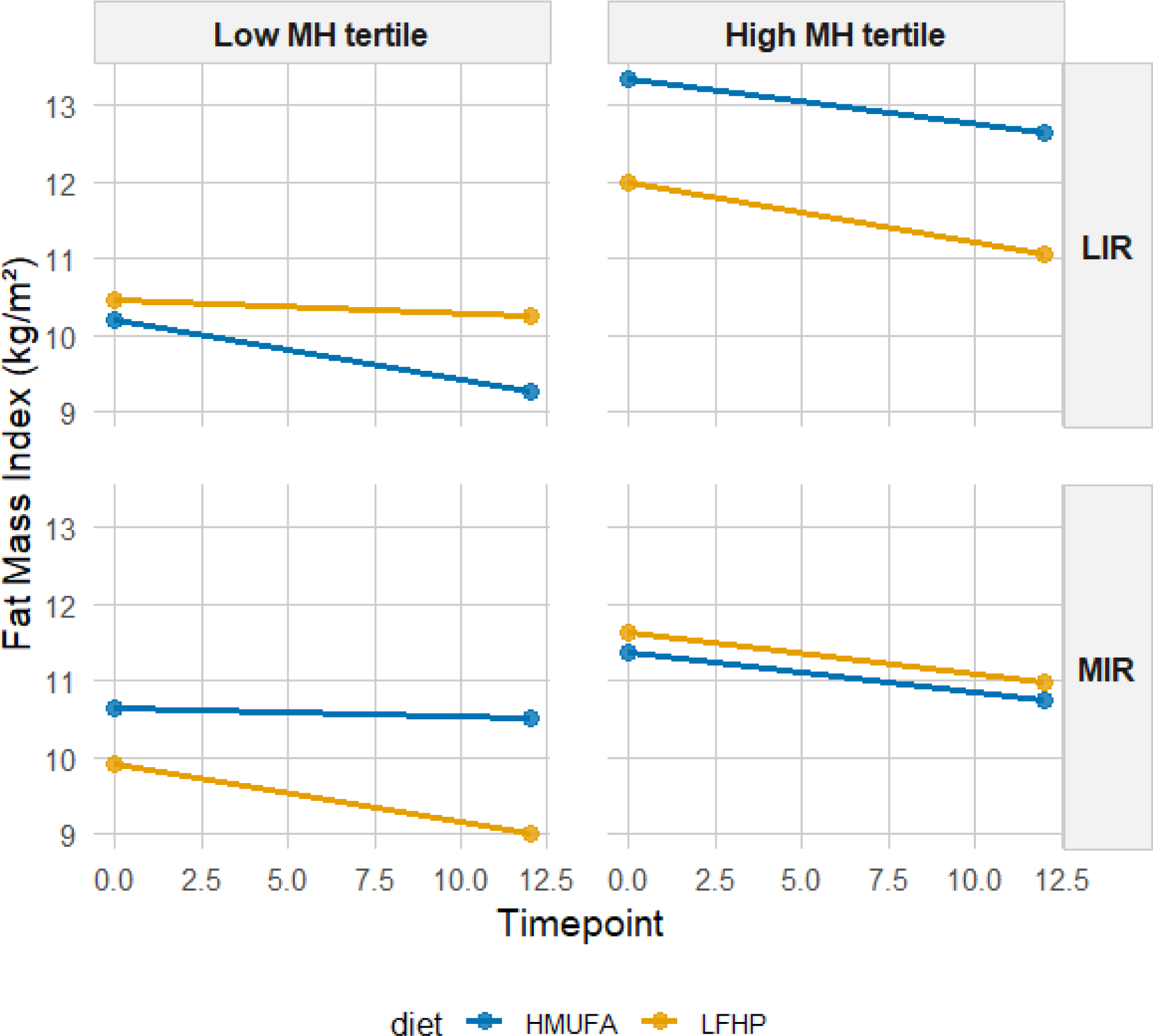
Estimated marginal means for fat mass index by IR metabotype and MH tertiles across diets. A 2×2 matrix shows estimated total fat percentage grouped by IR metabotype (MIR vs. LIR) and MH tertiles (high vs. low). The low-fat. high-protein. high-fiber (LFHP) diet is in yellow and the high-monounsaturated fat (HMUFA) diet in blue. LIR.Low (n=28). LIR.High (n=19). MIR.Low (n=34). MIR.High (n=36).

The LIR.LowMetaboHealth group experienced significant reductions in android fat on the HMUFA diet, while the LFHP diet showed no change. The LIR.HighMetaboHealth group showed reductions with both diets. In contrast, MIR.LowMetaboHealth had an increase in android fat on the HMUFA diet but a moderate fat loss on the LFHP diet, while participants in MIR.HighMetaboHealth benefited from reductions across both diets. For gynoid fat, LIR.LowMetaboHealth decreased on HMUFA but increased on LFHP, whereas the LIR.HighMetaboHealth group lost gynoid fat modestly with both diets. MIR.LowMetaboHealth maintained stable levels on LFHP but experienced an increase in HMUFA, and MIR.HighMetaboHealth saw slight losses regardless of diet. In terms of total fat percentage, LIR.LowMetaboHealth showed decreases in the HMUFA diet and a slight reduction in LFHP, with the LIR.HighMetaboHealth group also decreased on LFHP. MIR.LowMetaboHealth had minor changes on both diets, while MIR.HighMetaboHealth demonstrated reductions from both dietary approaches. Finally, regarding fat mass index, LIR.LowMetaboHealth benefited from the HMUFA diet and the LIR.HighMetaboHealth group showed improvements from both diets. The MIR.LowMetaboHealth group found the LFHP diet slightly more effective, while MIR.HighMetaboHealth experienced similar enhancements from both dietary interventions.

In summary, the results show that individuals in high MetaboHealth tertiles (those with poorer immune-metabolic health status) experienced reductions in android, gynoid, and total fat percentage and fat mass index regardless of metabotype and diet. At low MetaboHealth tertiles (those with relatively better immune-metabolic health status) the HMUFA diet largely benefitted individuals with the LIR metabotype, while the LFHP reduced android and gynoid fat in the MIR group.

## DISCUSSION

This study aimed to understand how personalized dietary interventions grouped by metabotype - specifically MIR and LIR - and MetaboHealth tertiles affect cardiometabolic health outcomes and body composition in a subset of 189 middle-aged and older adults that participated in the PERSON study (15). Stratifying for MetaboHealth did not refine the primary outcomes of the study. Our findings did show, however, that individuals classified in the higher MetaboHealth score tertile (reflecting poorer immune-metabolic health) demonstrate fat loss across both dietary regimens, while individuals in the lower MetaboHealth tertiles (reflecting better immune-metabolic health) exhibit improvement in android-upper-body and abdominal fat-and gynoid-lower-body fat-percentages when on the LFHP diet and expressing the MIR phenotype or when on the HMUFA diet and expressing the LIR phenotype. This study emphasizes that precision nutrition strategies can be refined by synergistic indicators of metabolic health status in managing body weight control effectively. Specifically, we observe a benefit of integrating baseline Metabohealth tertiles to understand the nuances of body composition changes, but not in cardiometabolic outcomes, across two guideline-abiding diets.

To the best of our knowledge, this is the first study to explore the interaction between by IR metabotypes and MetaboHealth tertiles to understand how middle-aged and older adults with tissue-specific IR respond to a personalized dietary intervention. Our findings indicate that the MetaboHealth score can serve as a valuable grouping tool in addition to IR metabotypes in future research (31,32). While initially set to improve cardiometabolic health parameters with insulin sensitivity as a primary outcome, this refined approach enhances our ability to predict which dietary approach - LFHP or HMUFA - will be more beneficial for middle-aged and older adults to elicit modest fat mass loss over three months, rather than changing IR in this time frame. This demonstrates that dietary interventions are not merely one-size-fits-all but should be informed by a comprehensive understanding of an individual’s cardiometabolic health profile (33) and an accompanying estimation of global immune metabolic health related to frailty and mortality as represented by MetaboHealth. Individuals in the highest MetaboHealth tertile experienced modest fat loss, irrespective of metabotype or diet intervention. This observation highlights how middle-aged and older adults with IR, unhealthier cardiometabolic health profiles, and similar higher fat mass percentages at baseline compared to those with healthier cardiometabolic health profiles, benefit from either of the two HMUFA or LFHP diets used in the PERSON study. In contrast, individuals with MIR in lower MetaboHealth tertiles benefitted significantly more from the LFHP diet, emphasizing the importance of global and tissue-specific IR related metabolic health status of individuals at baseline (34,35). Our results confirm the previous research within the PERSON study cohort, which detected benefits of the LFHP diet for the MIR group and the HMUFA diet for the LIR group when looking at cardiometabolic health outcomes, including measures of insulin sensitivity, triglycerides, and CRP (15). However, no differential changes in body composition were detected when participants were only grouped by metabotype (15). The present study therefore highlights the benefit of extending the grouping with the MetaboHealth score tertiles at baseline, representing other aspects of metabolic health, to detect changes in body composition, but not necessarily in cardiometabolic outcomes, across personalized dietary interventions.

The mechanisms underlying the observed responses to dietary interventions likely involve key interactions between dietary composition, insulin signaling pathways, and changes in body composition (36–38). In individuals with LIR, the HMUFA diet, rich in monounsaturated fatty acids, may provide specific benefits by improving hepatic lipid metabolism and reducing inflammation (36,37). Previous research suggests that monounsaturated fatty acids may usher body fat loss specifically via the activation of AMP-activated protein kinase signaling (38,39). Overall, monounsaturated fatty acids enhance insulin sensitivity, optimize lipid profiles, and decrease the accumulation of harmful lipotoxic intermediates that can disrupt metabolic function (40–42). Additionally, individuals with MIR following the LFHP diet may ensure higher protein intake, positively influencing insulin signaling and likely promoting the activation of pathways involved in glucose metabolism and muscle maintenance (43,44). This interplay may ensure effective nutrient utilization, facilitates energy balance, and promotes fat oxidation while preventing excessive fat storage (45,46). By incorporating tailored dietary strategies, it may be possible to optimize health outcomes, possibly through these complex mechanistic pathways. Interestingly, these findings contrast with the results from the CORDIOPREV study, where LIR individuals benefitted from the LFHP diet and the MIR individuals from HMUFA in terms of cardiometabolic indicators (47). While the latter study did not include DXA measurements, the study did detect changes after two years in anthropometric BMI and waist circumference measurements in the LIR group when provided with the LFHP diet, but not when provided with the Mediterranean diet - akin to the HMUFA in the PERSON study (48). However, previous research on the PERSON study and our study demonstrates the exact opposite, where LIR participants largely benefit both in cardiometabolic health and body composition outcomes (15). The difference may be due to the CORDIOPREV cohort being largely composed of type 2 diabetes, while the PERSON study excluded those with type 2 diabetes. Future studies should investigate different groups of metabolic status in IR metabotypes with complete analyses of body composition to understand the distribution of fat loss, and whether lean mass is preserved, across dietary interventions.

While our study demonstrated minor android, gynoid, total fat percentage, and fat mass indices loss with both dietary interventions, we did not observe improvements in the MetaboHealth score upon the intervention. It is important to note that the score was initially created as a risk indicator to predict long-term (5-10 year) mortality with the ability to detect an older individual’s global long-term health status determined by muscle, cognitive health and frailty, which the score indicates (18,20). This study shows that the MetaboHealth score as an outcome may not be particularly sensitive to short-term dietary changes and accompanying metabolic health improvements in IR individuals in this cohort. We, therefore, conclude that, although MetaboHealth effectively groups participants in tertiles at baseline, it may not adequately capture the nuances of cardiometabolic health changes in IR by short-term dietary interventions. For the MetaboHealth score to change, especially VLDL and HDL particle size, the ratio of polyunsaturated fatty acids to total fatty acids (PUFA %), glucose, amino acids, albumin, and a low grade inflammation markers glycoprotein acetyls would have to change, which does not seems to be the case by the HMUFA or LFHP diets. Other short-term interventions of individuals above 60 years of age, including ones in which diet and physical exercise are combined, are being explored in parallel showing that especially interventions that change low-grade inflammation appear to be recorded by the MetaboHealth score. Although it is not necessarily the most sensitive monitor for such effects MetaboHealth does show improvement of global health (49,50).

The strengths of this study include its robust design as part of a randomized controlled trial, the grouping of participants based on detailed metabolic profiles, and the use of well-validated, such as DXA for body composition assessment, and high-throughput targeted metabolomic analysis. The grouping into MetaboHealth score tertiles allows for a more nuanced understanding of body composition cardiometabolic health and its impact on dietary responses. However, there are limitations to consider. The relatively short duration of the dietary interventions may not capture long-term metabolic adaptations. Future studies should investigate the sustainability of these dietary approaches over extended periods. Additionally, the reliance on specific dietary regimens may not encompass the broader spectrum of dietary patterns beneficial for diverse populations, limiting the generalizability of our findings to other dietary patterns. Future research should explore the MetaboHealth score and other novel metabolomics or proteomics biomarkers or composite scores to monitor the response to lifestyle interventions. Moreover, given that these results were fundamentally post-hoc analyses, it may be plausible to imagine that this study was underpowered as evidenced by the small groups.

In conclusion, our findings suggest that personalized dietary strategies for middle-aged and older adults with insulin resistance can be more effective when considering both the specific metabolic phenotype and the broader MetaboHealth score defined in tertiles at baseline. This may indicate that adding global markers of immune-metabolic health to disease-specific ones when investigating responses of adults to interventions may provide outcome benefits. By integrating MetaboHealth score grouping into personalized dietary approaches, we offer valuable insights into optimizing body-compositional health outcomes in insulin-resistant individuals. Future interventions using a combination of personalized dietary regimens offer a comprehensive approach to managing IR and improving overall physical and metabolic health.

## Data Availability

All data produced in the present study are available upon reasonable request to the authors from WUR and MUMC.

## ACKNOWLEDGMENTS

The authors wish to acknowledge the services of the PERSON study, the contributing research centres, and all the study participants. We thank the PERSON study team at both WUR and MUMC for their invaluable contribution to the execution of the study.

This work is funded by the VOILA Consortium (ZonMw 457001001), which had no role in the design and conduct of the study; collection, management, analysis, and interpretation of the data; and preparation, review, or approval of the manuscript.

---

**FIGURE S1. FIGURE S4.**
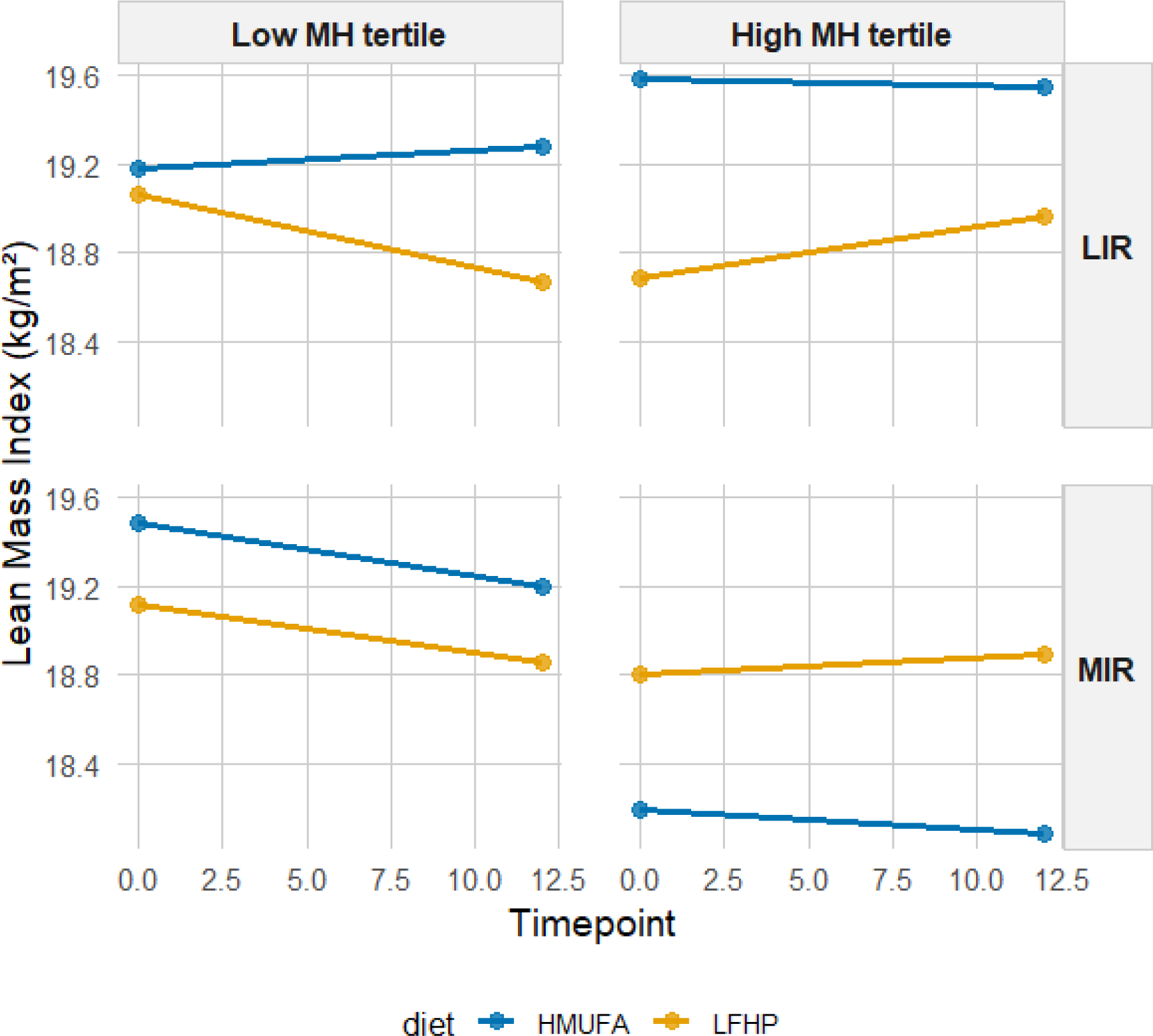
Estimated marginal means for lean mass index by IR metabotype and MH tertiles across diets. A 2×2 matrix shows estimated total fat percentage grouped by IR metabotype (MIR vs. LIR) and MH tertiles (high vs. low). The low-fat. high-protein. high-fiber (LFHP) diet is in yellow and the high-monounsaturated fat (HMUFA) diet in blue. LIR.Low (n=28). LIR.High (n=19). MIR.Low (n=34). MIR.High (n=36).

**FIGURE S2.**
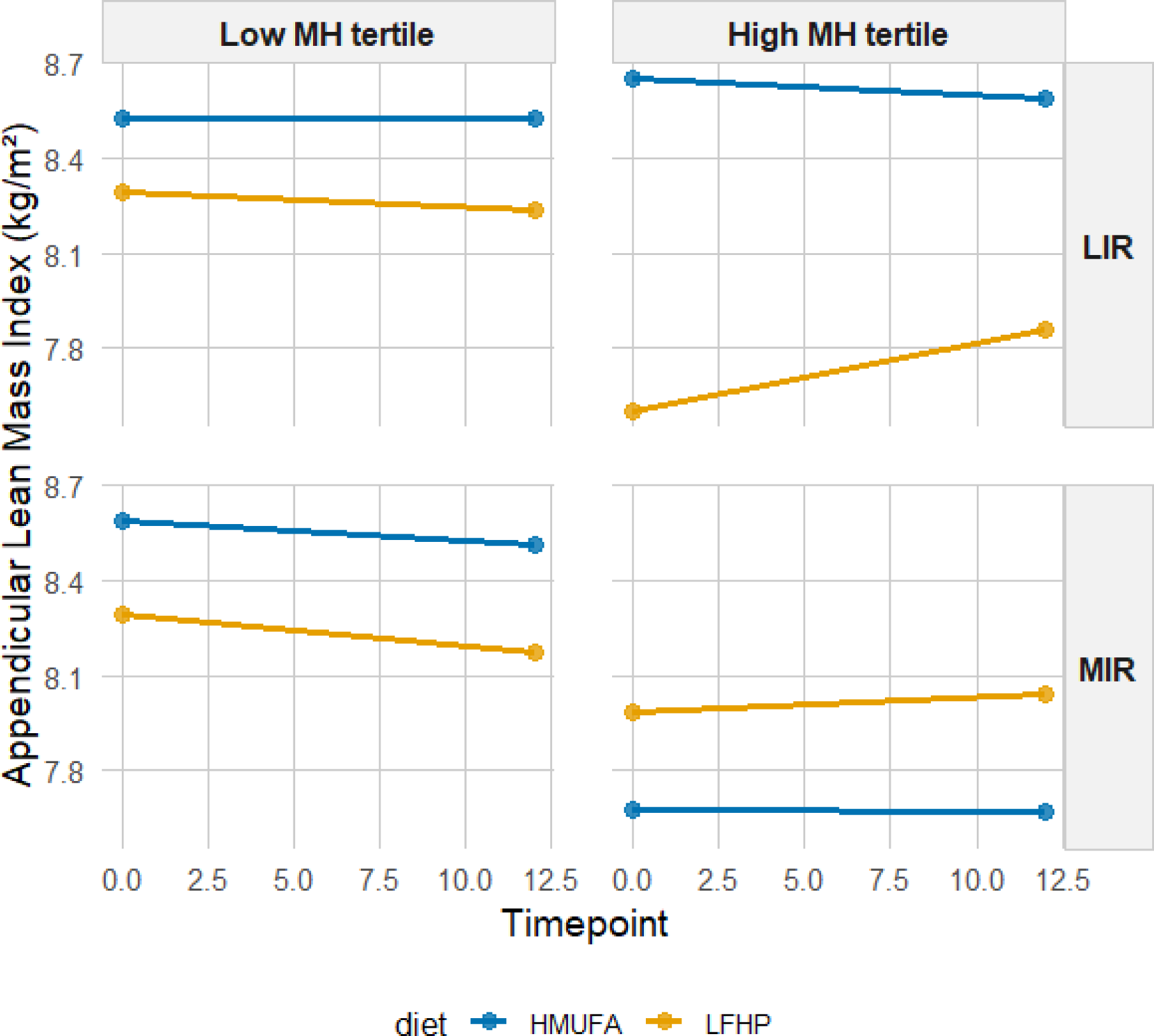
Estimated marginal means for appendicular lean mass index by IR metabotype and MH tertiles across diets. A 2×2 matrix shows estimated total fat percentage grouped by IR metabotype (MIR vs. LIR) and MH tertiles (high vs. low). The low-fat. high-protein. high-fiber (LFHP) diet is in yellow and the high-monounsaturated fat (HMUFA) diet in blue. LIR.Low (n=28). LIR.High (n=19). MIR.Low (n=34). MIR.High (n=36).

**FIGURE S3.**
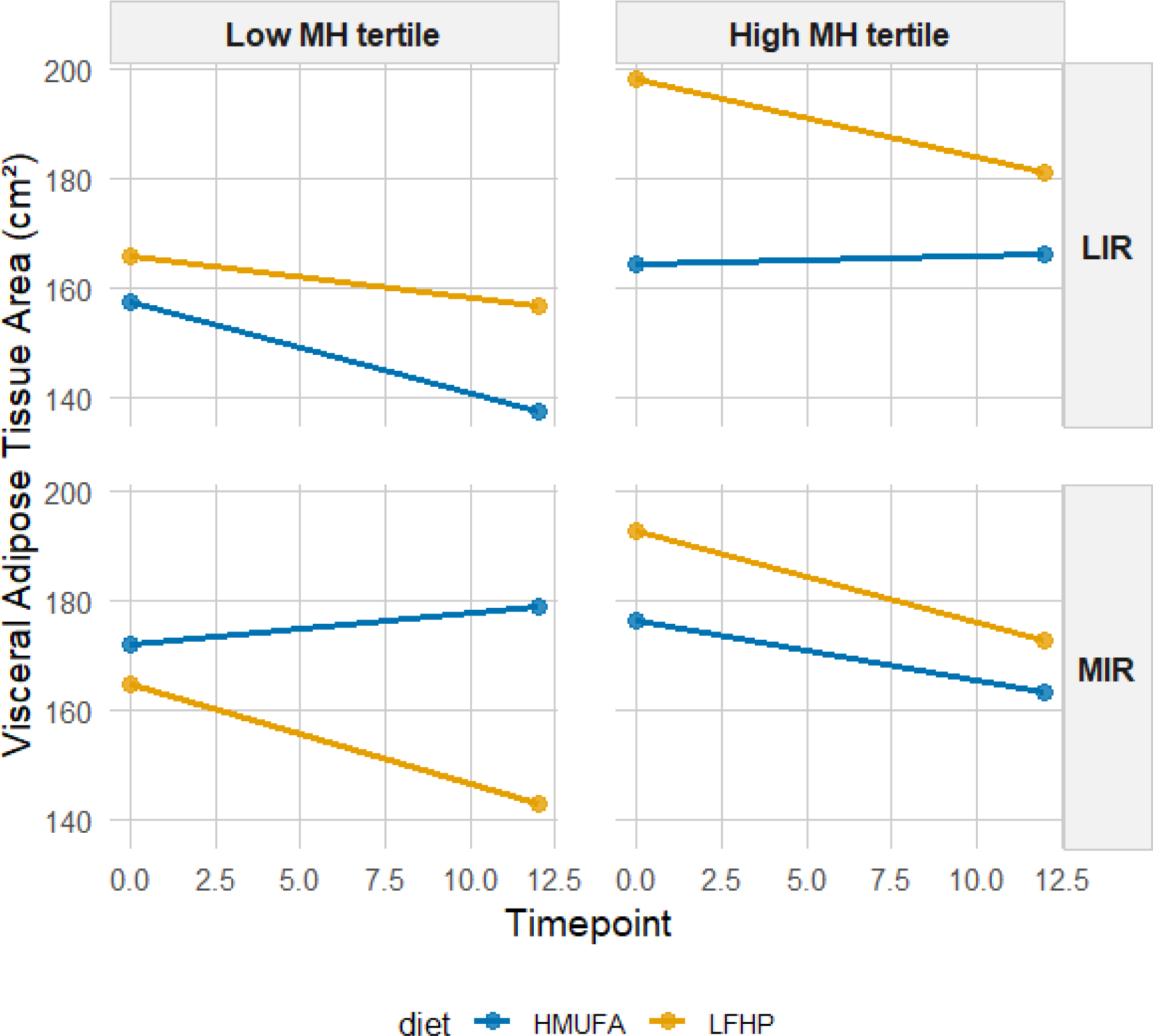
Estimated marginal means for VAT by IR metabotype and MH tertiles across diets. A 2×2 matrix shows estimated total fat percentage grouped by IR metabotype (MIR vs. LIR) and MH tertiles (high vs. low). The low-fat. high-protein. high-fiber (LFHP) diet is in yellow and the high-monounsaturated fat (HMUFA) diet in blue. LIR.Low (n=28). LIR.High (n=19). MIR.Low (n=34). MIR.High (n=36).

**FIGURE S4.**
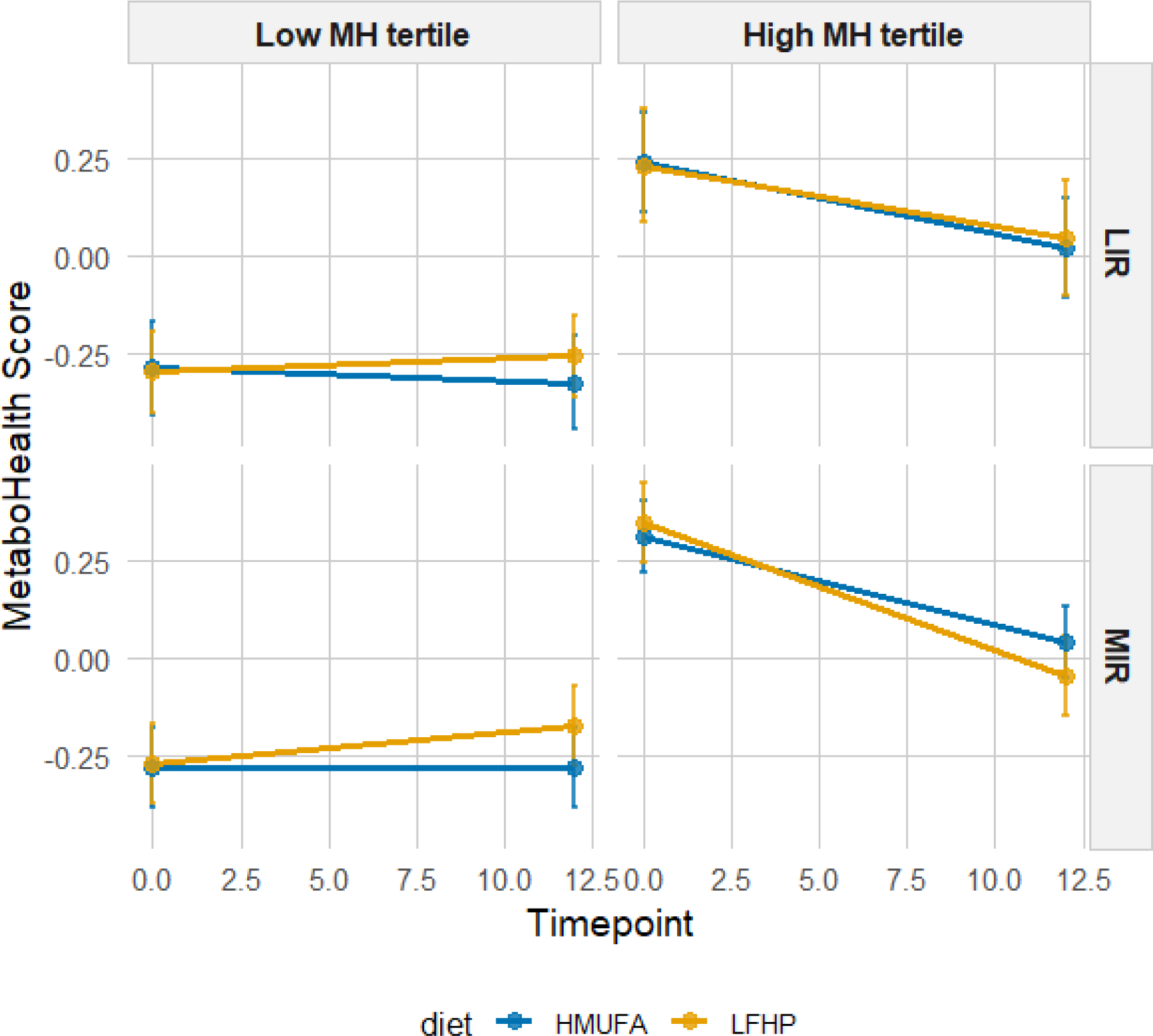
Estimated marginal means for MetaboHealth score by IR metabotype and MH tertiles across diets. A 2×2 matrix shows estimated total fat percentage grouped by IR metabotype (MIR vs. LIR) and MH tertiles (high vs. low). The low-fat. high-protein high-fiber (LFHP) diet is in yellow and the high-monounsaturated fat (HMUFA) diet in blue. LIR.Low (n=28). LIR.High (n=19). MIR.Low (n=34). MIR.High (n=36).

**FIGURE S5.**
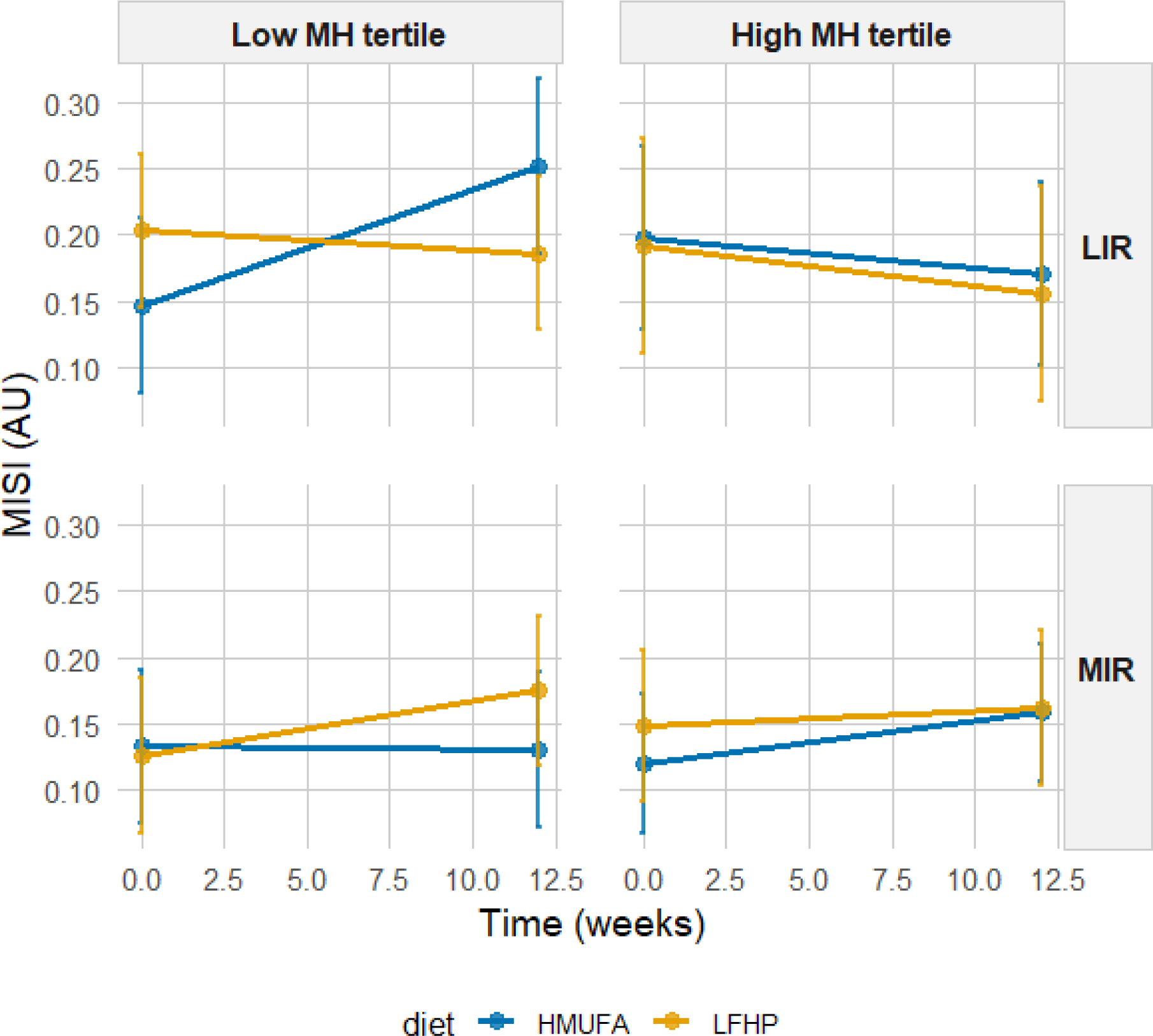
Estimated marginal means for MISI by IR metabotype and MH tertiles across diets. A 2×2 matrix shows estimated total fat percentage grouped by IR metabotype (MIR vs. LIR) and MH tertiles (high vs. low). The low-fat. high-protein high-fiber (LFHP) diet is in yellow and the high-monounsaturated fat (HMUFA) diet in blue. LIR.Low (n=28). LIR.High (n=19). MIR.Low (n=34). MIR.High (n=36).

**FIGURE S6.**
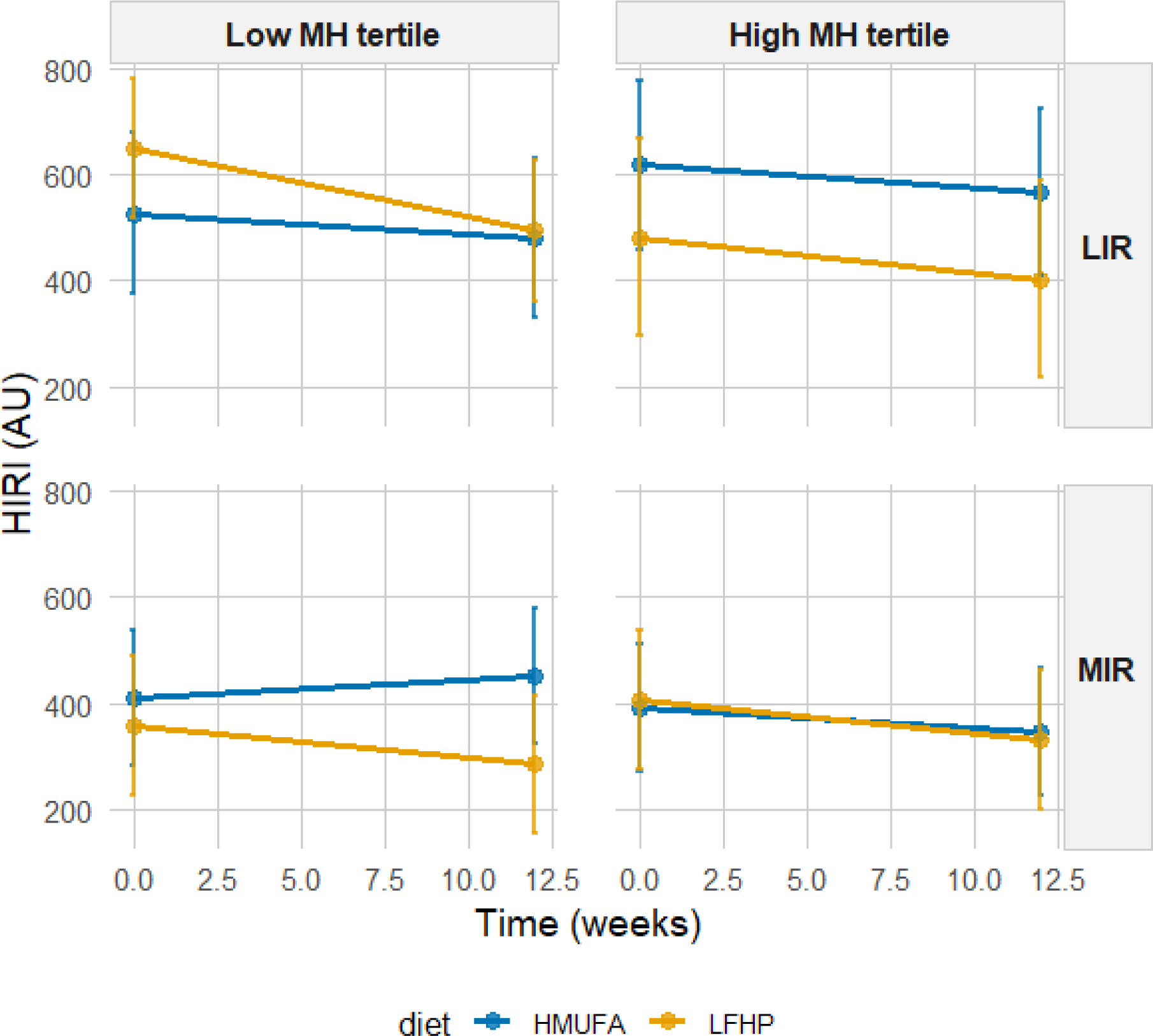
Estimated marginal means for HIRI by IR metabotype and MH tertiles across diets. A 2×2 matrix shows estimated total fat percentage grouped by IR metabotype (MIR vs. LIR) and MH tertiles (high vs. low). The low-fat. high-protein high-fiber (LFHP) diet is in yellow and the high-monounsaturated fat (HMUFA) diet in blue. LIR.Low (n=28). LIR.High (n=19). MIR.Low (n=34). MIR.High (n=36).

**FIGURE S7.**
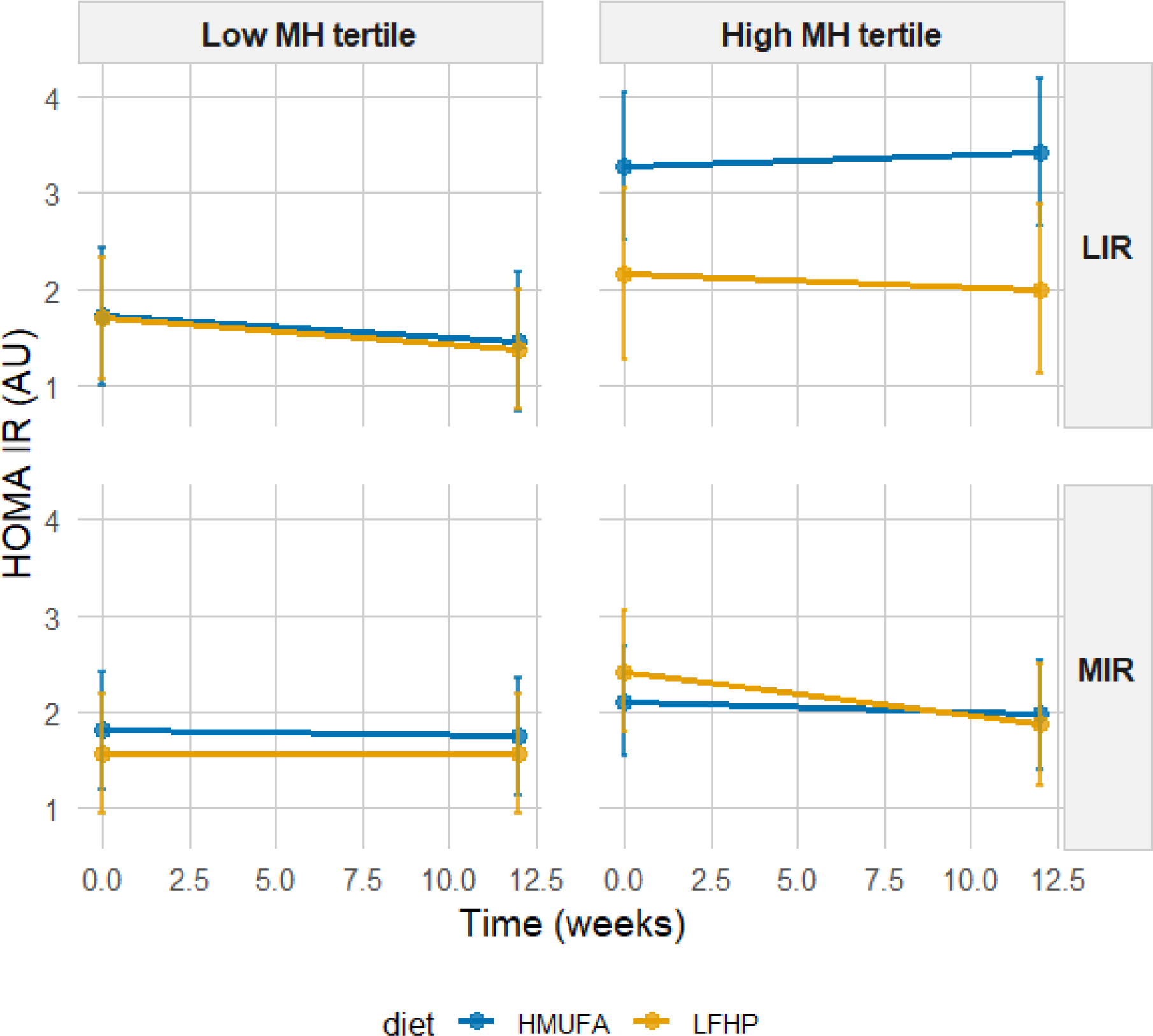
Estimated marginal means for HOMA-IR by IR metabotype and MH tertiles across diets. A 2×2 matrix shows estimated total fat percentage grouped by IR metabotype (MIR vs. LIR) and MH tertiles (high vs. low). The low-fat. high-protein high-fiber (LFHP) diet is in yellow and the high-monounsaturated fat (HMUFA) diet in blue. LIR.Low (n=28). LIR.High (n=19). MIR.Low (n=34). MIR.High (n=36).

**FIGURE S8.**
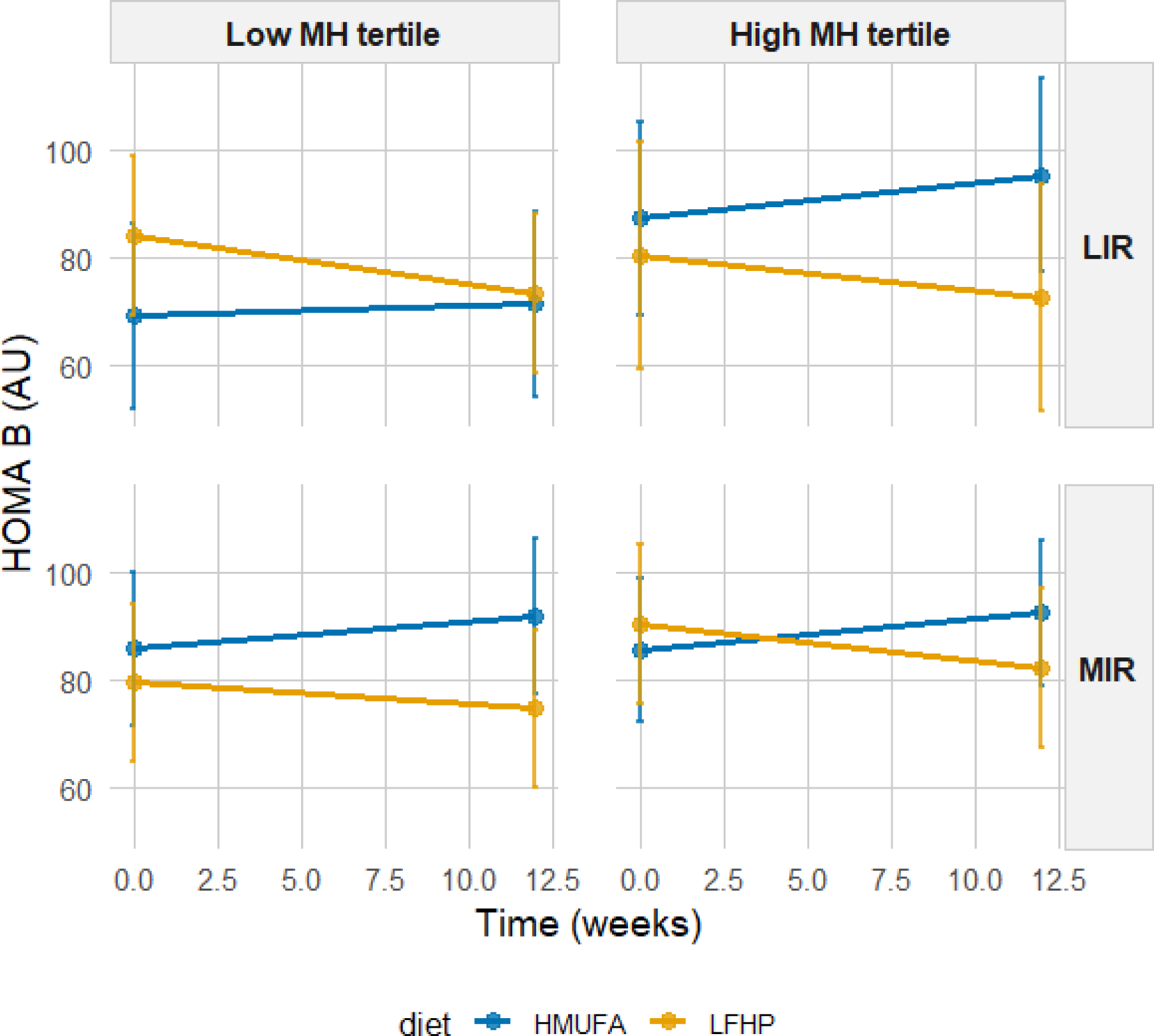
Estimated marginal means for HOMA-B by IR metabotype and MH tertiles across diets. A 2×2 matrix shows estimated total fat percentage grouped by IR metabotype (MIR vs. LIR) and MH tertiles (high vs. low). The low-fat. high-protein high-fiber (LFHP) diet is in yellow and the high-monounsaturated fat (HMUFA) diet in blue. LIR.Low (n=28). LIR.High (n=19). MIR.Low (n=34). MIR.High (n=36).

**FIGURE S9.**
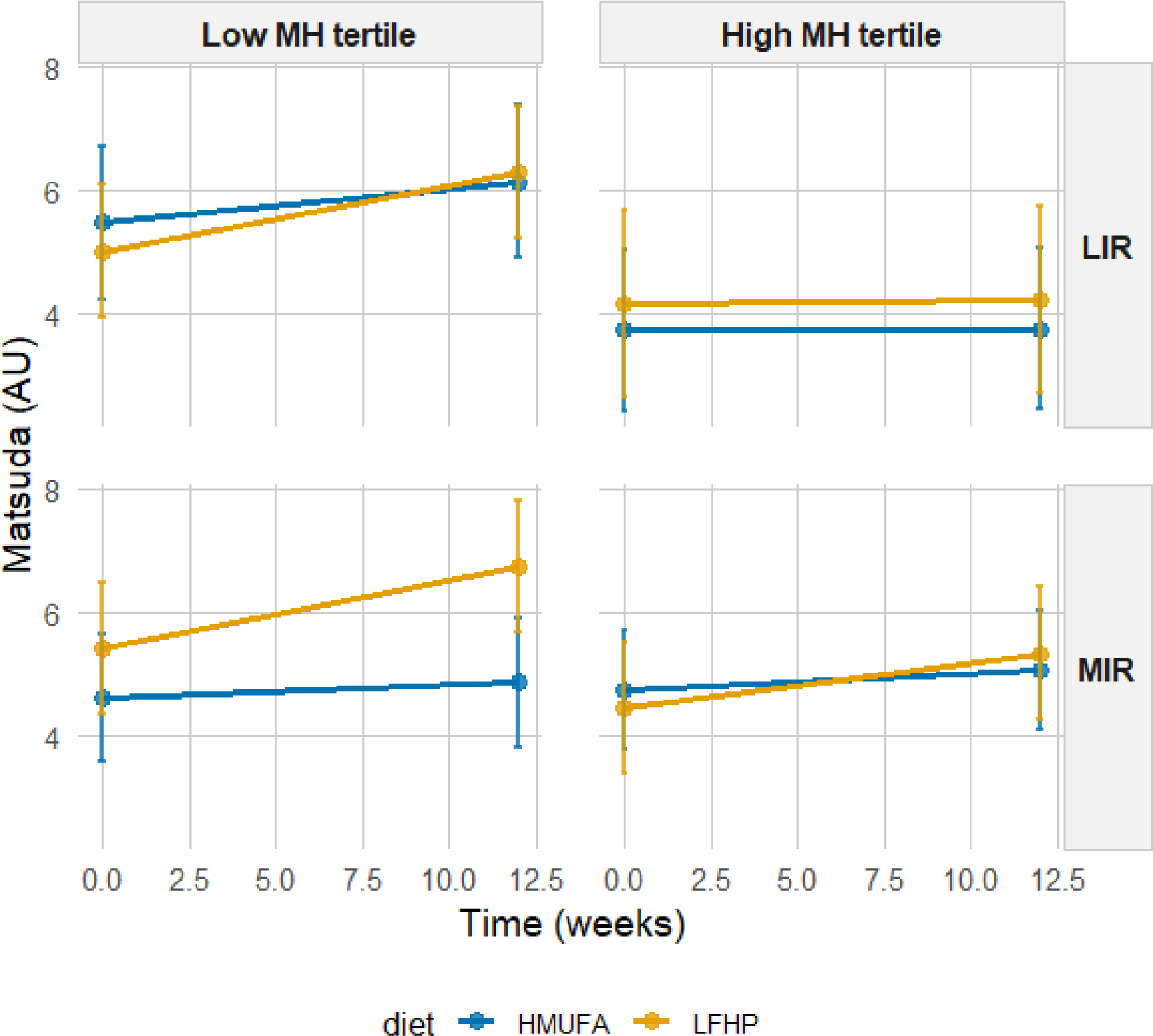
Estimated marginal means for Matsuda Index by IR metabotype and MH tertiles across diets. A 2×2 matrix shows estimated total fat percentage grouped by IR metabotype (MIR vs. LIR) and MH tertiles (high vs. low). The low-fat. high-protein high-fiber (LFHP) diet is in yellow and the high-monounsaturated fat (HMUFA) diet in blue. LIR.Low (n=28). LIR.High (n=19). MIR.Low (n=34). MIR.High (n=36).

**FIGURE S10.**
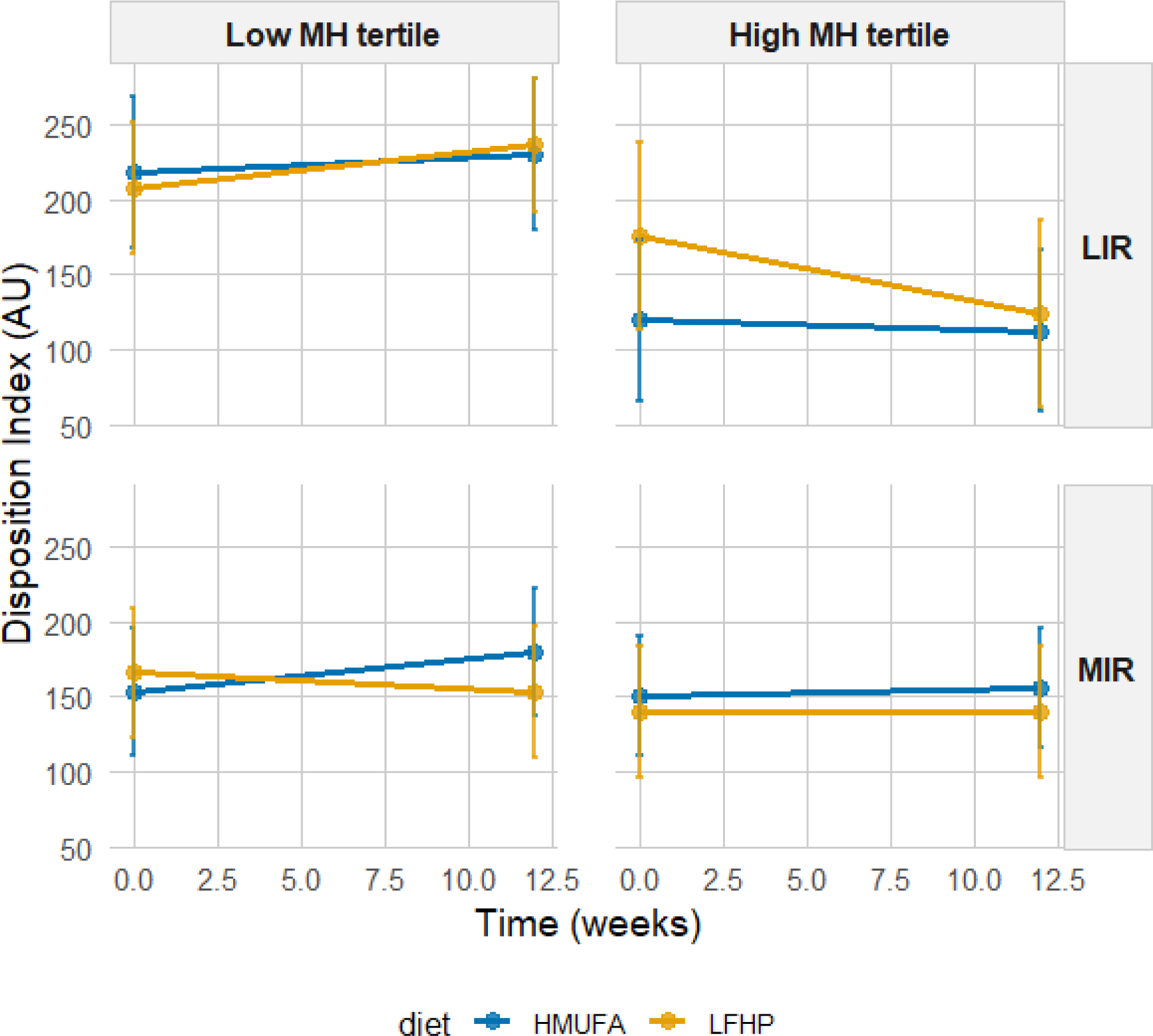
Estimated marginal means for Disposition Index by IR metabotype and MH tertiles across diets. A 2×2 matrix shows estimated total fat percentage grouped by IR metabotype (MIR vs. LIR) and MH tertiles (high vs. low). The low-fat. high-protein high-fiber (LFHP) diet is in yellow and the high-monounsaturated fat (HMUFA) diet in blue. LIR.Low (n=28). LIR.High (n=19). MIR.Low (n=34). MIR.High (n=36).

**FIGURE S11.**
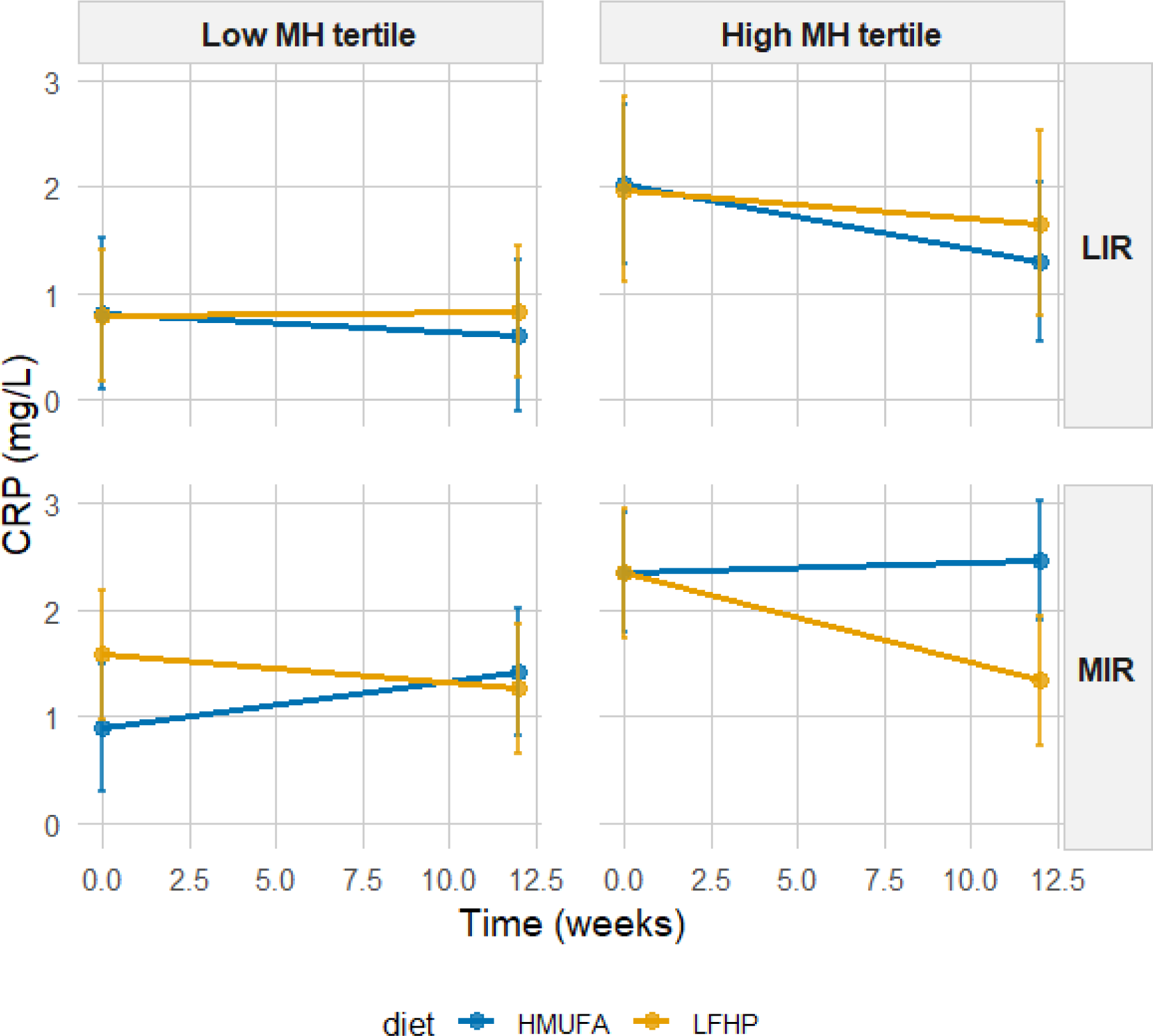
Estimated marginal means for CRP by IR metabotype and MH tertiles across diets. A 2×2 matrix shows estimated total fat percentage grouped by IR metabotype (MIR vs. LIR) and MH tertiles (high vs. low). The low-fat. high-protein high-fiber (LFHP) diet is in yellow and the high-monounsaturated fat (HMUFA) diet in blue. LIR.Low (n=28). LIR.High (n=19). MIR.Low (n=34). MIR.High (n=36).

**FIGURE S12.**
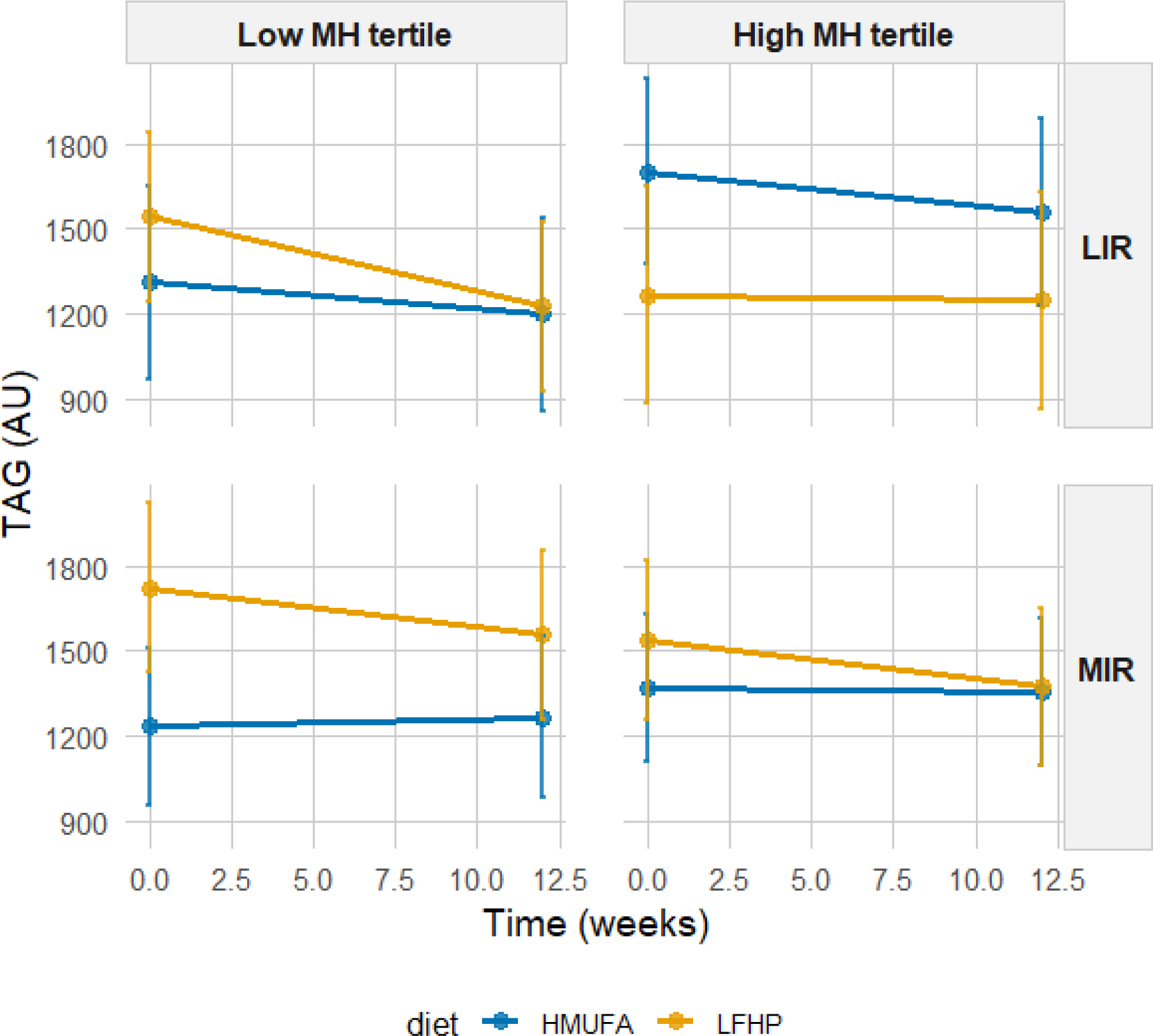
Estimated marginal means for TAG by IR metabotype and MH tertiles across diets. A 2×2 matrix shows estimated total fat percentage grouped by IR metabotype (MIR vs. LIR) and MH tertiles (high vs. low). The low-fat. high-protein high-fiber (LFHP) diet is in yellow and the high-monounsaturated fat (HMUFA) diet in blue. LIR.Low (n=28). LIR.High (n=19). MIR.Low (n=34). MIR.High (n=36).

## ABBREVIATIONS

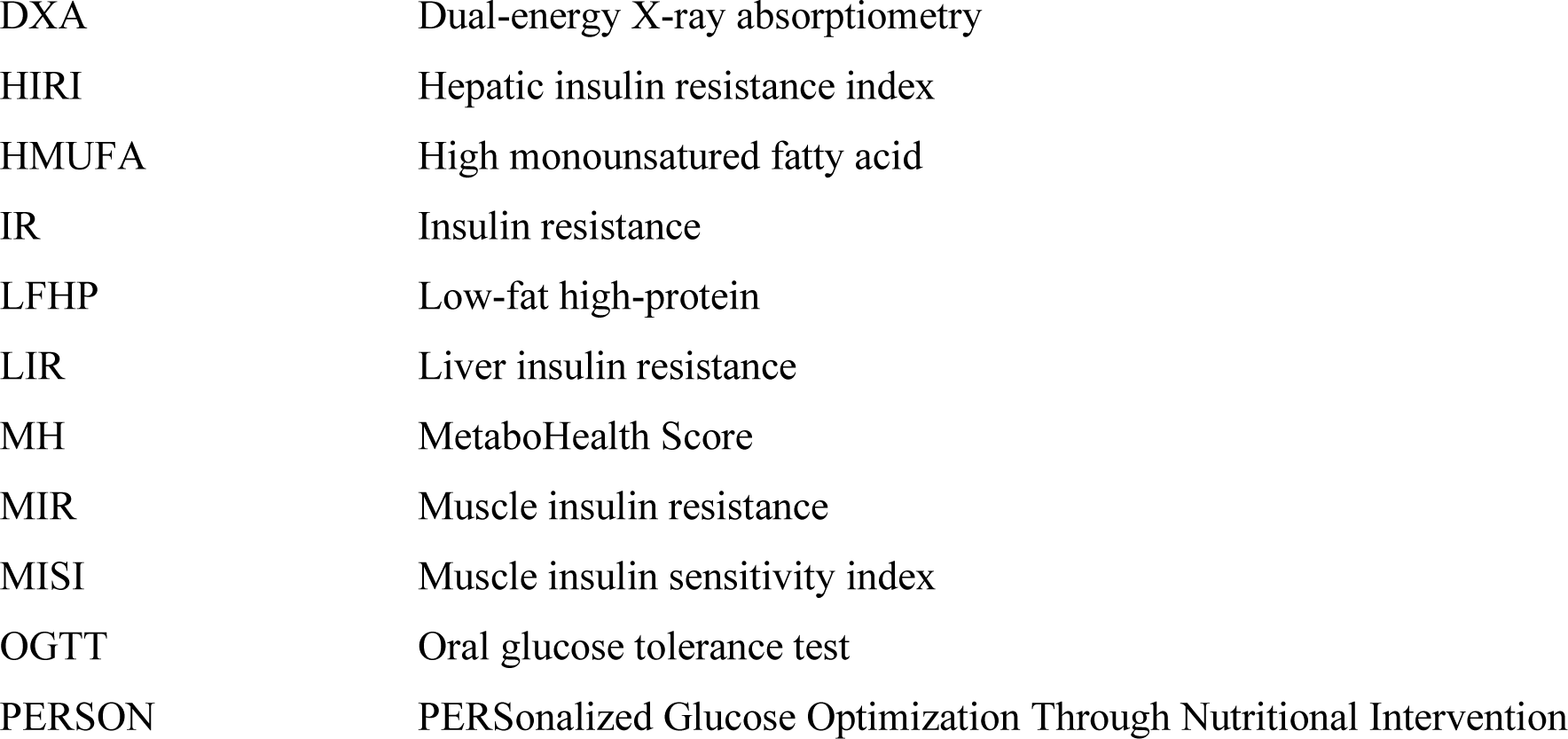

